# Phenotyping of acute and persistent COVID-19 features in the outpatient setting: exploratory analysis of an international cross-sectional online survey

**DOI:** 10.1101/2021.08.05.21261677

**Authors:** Sabina Sahanic, Piotr Tymoszuk, Dietmar Ausserhofer, Verena Rass, Alex Pizzini, Goetz Nordmeyer, Katharina Hüfner, Katharina Kurz, Paulina Maria Weber, Thomas Sonnweber, Anna Boehm, Magdalena Aichner, Katharina Cima, Barbara Boeckle, Bernhard Holzner, Gerhard Rumpold, Christoph Puelacher, Stefan Kiechl, Andreas Huber, Christian J. Wiedermann, Barbara Sperner-Unterweger, Ivan Tancevski, Rosa Bellmann-Weiler, Herbert Bachler, Giuliano Piccoliori, Raimund Helbok, Guenter Weiss, Judith Loeffler-Ragg

## Abstract

**BACKGROUND:** Long COVID, defined as presence of COVID-19 related symptoms 28 days or more after the onset of acute SARS-CoV-2 infection, is an emerging challenge to healthcare systems. The objective of this study was to phenotype recovery trajectories of non-hospitalized COVID-19 individuals.

**METHODS:** We performed an international, multi-center, exploratory online survey study on demographics, comorbidities, COVID-19 symptoms and recovery status of non-hospitalized SARS-CoV-2 infected adults (Austria: n=1157), and Italy: n= 893).

**RESULTS:** Working age subjects (Austria median: 43 yrs (IQR: 31 – 53), Italy: 45 yrs (IQR: 35 – 55)) and females (65.1% and 68.3%) predominated the study cohorts. Course of acute COVID-19 was characterized by a high symptom burden (median 13 (IQR: 9 – 18) and 13 (7 – 18) out of 44 features queried), a 47.6 – 49.3% rate of symptom persistence beyond 28 days and 20.9 – 31.9% relapse rate. By cluster analysis, two acute symptom phenotypes could be discerned: the non-specific infection phenotype and the multi-organ phenotype (MOP), the latter encompassing multiple neurological, cardiopulmonary, gastrointestinal and dermatological features. Clustering of long COVID subjects yielded three distinct subgroups, with a subset of 48.7 – 55 % long COVID individuals particularly affected by post-acute MOP symptoms. The number and presence of specific acute MOP symptoms and pre-existing multi-morbidity was linked to elevated risk of long COVID.

**CONCLUSION:** The consistent findings of two independent cohorts further delineate patterns of acute and post-acute COVID-19 and emphasize the importance of symptom phenotyping of home-isolated COVID-19 patients to predict protracted convalescence and to allocate medical resources.

**Key Points:** *Question:* Which acute symptom patterns of acute COVID-19 are associated with prolonged symptom persistence, symptom relapse or physical performance impairment?

*Findings:* In this multicenter international comparative survey study on non-hospitalized SARS-CoV-2 infected adults (Austria: n = 1157, Italy: n = 893) we identified distinct and reproducible phenotypes of acute and persistent features. Acute multi-organ symptoms including neurological and cardiopulmonary manifestations are linked to elevated risk of long COVID.

*Meaning:* These findings suggest to employ symptom phenotyping of home-isolated COVID-19 patients to predict protracted convalescence and to allocate medical resources.

## Introduction

Coronavirus disease 2019 (COVID-19) displays a broad clinical spectrum ranging from asymptomatic to fatal courses of infection(1) and a large variability of symptom duration beyond acute illness.

The implications and consequences of such long-lasting, in many cases relapsing complaints for more than four weeks, often described as ‘long COVID’, are a growing health concern and a new burden to the health care systems.(2) For the sake of medical resource allocation, the terms ‘ongoing symptomatic COVID-19’ for persistence of post-infectious symptoms for 4 – 12 weeks and ‘post-COVID-19 syndrome’ for symptoms lasting for 12 weeks or longer were introduced.(3)

Long-lasting dyspnoea and fatigue are well characterized in hospitalized COVID-19 patients. Little is known about the clinical phenotype of outpatients, who constitute the majority of symptomatic COVID-19 cases. (4–6) Recent evidence suggests, that also this subset, commonly classified as mild COVID-19, experiences prolonged symptoms including chronic cough, shortness of breath, chest tightness, cognitive dysfunction, and fatigue.(7–10) For this group, identification of subjects at risk of long COVID is urgently needed to effectively allocate healthcare resources.

With this international multi-center online survey of non-hospitalized COVID-19 patients, we sought to describe prevalence and patterns of acute and persistent manifestations and identify key factors impacting the presence and relapse of persistent symptoms and major physical impairment.

## Methods

### Study design and participants

The multi-center study ‘Health after COVID-19 in Tyrol’ (ClinicalTrials.gov: NCT04661462) was conducted in the neighboring European regions Tyrol (Austria) and South Tyrol (Italy) between the 30^th^September 2020 and 5^th^ July 2021 as an anonymized online survey.(11) The participants were invited via public media calls (Tyrol and South Tyrol) and by general practitioners (South Tyrol). The inclusion criteria encompassed confirmed SARS-CoV2 infection (PCR or seropositivity), Tyrol/South Tyrol residence and age ≥ 16 (Tyrol) or ≥18 years (South Tyrol). Exclusion criteria from further analysis were hospitalization because of COVID-19 and survey completion < 28 days after initial diagnosis of the infection (**Figure 1**).

**Figure 1.**
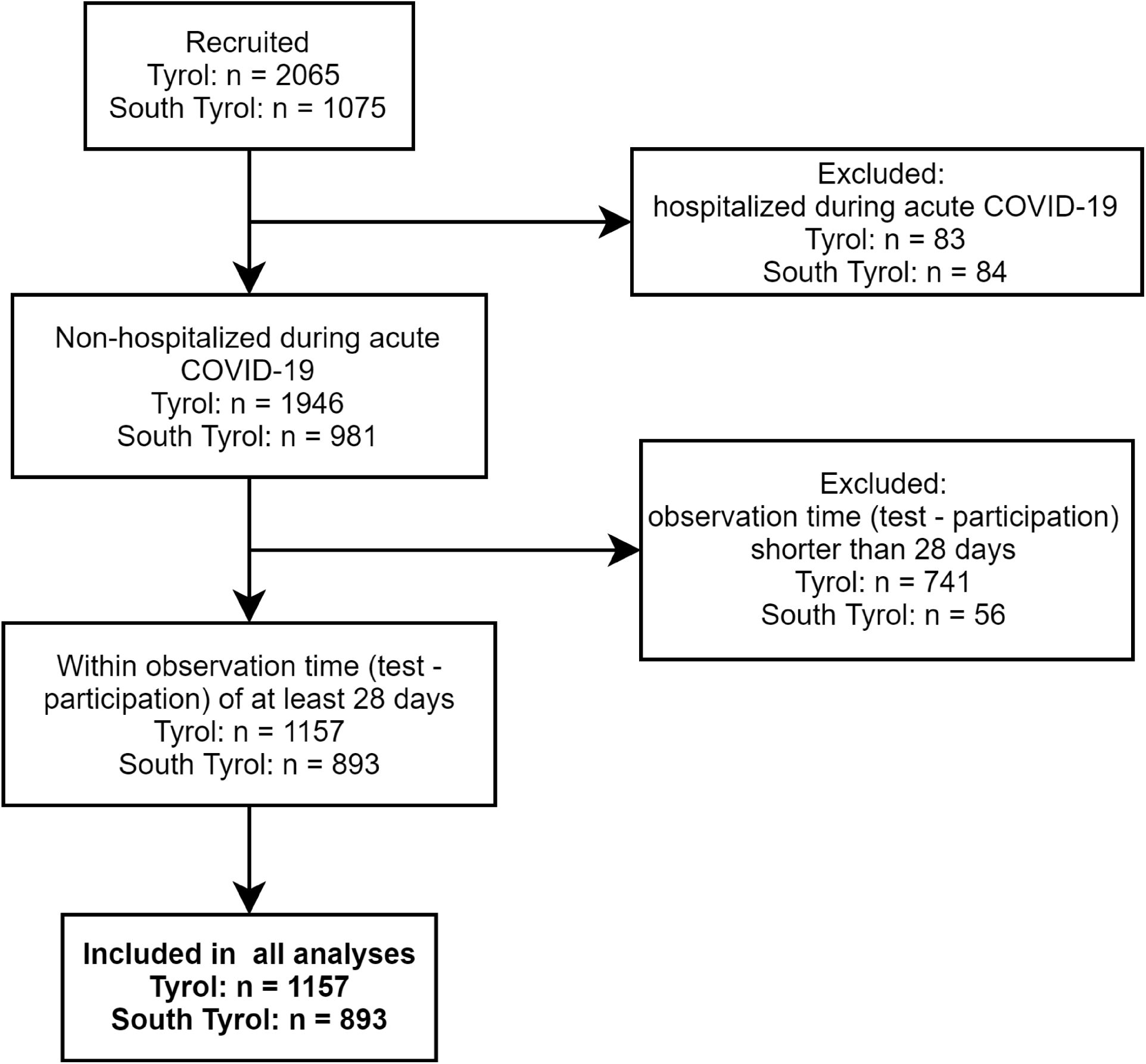
CONSORT flow diagram for the study population.

This study was approved by the institutional review boards of the Medical University of Innsbruck (Austria) (approval number: 1257/2020) and the South Tyrol Province (Italy) (0150701). Each participant gave a digital informed consent at the survey start.

### Measures

Data on demographics, socioeconomic status, comorbidities, smoking, daily medication relevant to SARS-CoV-2 infection, course and perception of acute SARS-CoV-2 infection, presence/duration of COVID-19-associated symptoms (44 items), symptom relapse and physical constitution during convalescence were recorded (**Supplementary Table S1**). The baseline cohort characteristics are shown in **Table 1 and 2**, the features of acute and post-acute COVID-19 in **Table 3**.

**Table 1.**
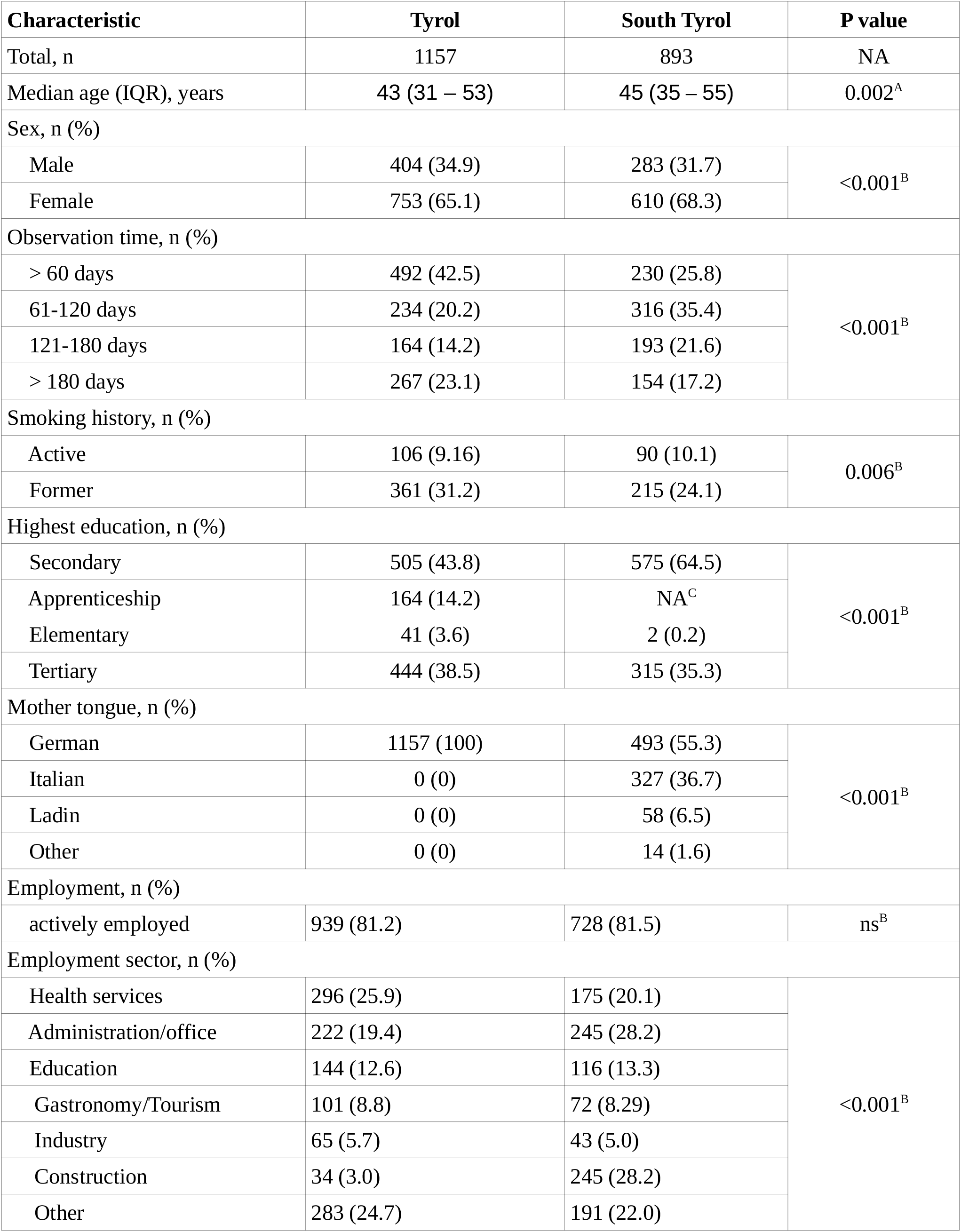
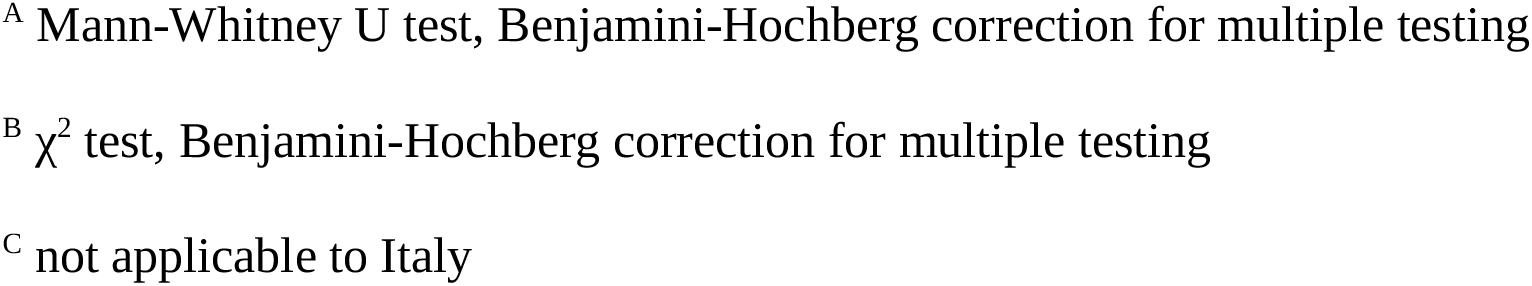
Sociodemographic characteristics of the study cohorts.

| Characteristic | Tyrol | South Tyrol | P value |
| --- | --- | --- | --- |
| Total, n | 1157 | 893 | NA |
| Median age (IQR), years | 43 (31 – 53) | 45 (35 – 55) | 0.002<sup>A</sup> |
| Sex, n (%) |  |  |  |
| Male | 404 (34.9) | 283 (31.7) | <0.001<sup>B</sup> |
| Female | 753 (65.1) | 610 (68.3) |  |
| Observation time, n (%) |  |  |  |
| > 60 days | 492 (42.5) | 230 (25.8) | <0.001<sup>B</sup> |
| 61-120 days | 234 (20.2) | 316 (35.4) |  |
| 121-180 days | 164 (14.2) | 193 (21.6) |  |
| > 180 days | 267 (23.1) | 154 (17.2) |  |
| Smoking history, n (%) |  |  |  |
| Active | 106 (9.16) | 90 (10.1) | 0.006<sup>B</sup> |
| Former | 361 (31.2) | 215 (24.1) |  |
| Highest education, n (%) |  |  |  |
| Secondary | 505 (43.8) | 575 (64.5) | <0.001<sup>B</sup> |
| Apprenticeship | 164 (14.2) | NA<sup>C</sup> |  |
| Elementary | 41 (3.6) | 2 (0.2) |  |
| Tertiary | 444 (38.5) | 315 (35.3) |  |
| Mother tongue, n (%) |  |  |  |
| German | 1157 (100) | 493 (55.3) | <0.001<sup>B</sup> |
| Italian | 0 (0) | 327 (36.7) |  |
| Ladin | 0 (0) | 58 (6.5) |  |
| Other | 0 (0) | 14 (1.6) |  |
| Employment, n (%) |  |  |  |
| actively employed | 939 (81.2) | 728 (81.5) | ns<sup>B</sup> |
| Employment sector, n (%) |  |  |  |
| Health services | 296 (25.9) | 175 (20.1) | <0.001<sup>B</sup> |
| Administration/office | 222 (19.4) | 245 (28.2) |  |
| Education | 144 (12.6) | 116 (13.3) |  |
| Gastronomy/Tourism | 101 (8.8) | 72 (8.29) |  |
| Industry | 65 (5.7) | 43 (5.0) |  |
| Construction | 34 (3.0) | 245 (28.2) |  |
| Other | 283 (24.7) | 191 (22.0) |  |

<sup>A</sup> Mann-Whitney U test, Benjamini-Hochberg correction for multiple testing
<sup>B</sup> $\chi^2$ test, Benjamini-Hochberg correction for multiple testing
<sup>C</sup> not applicable to Italy

**Table 2.**
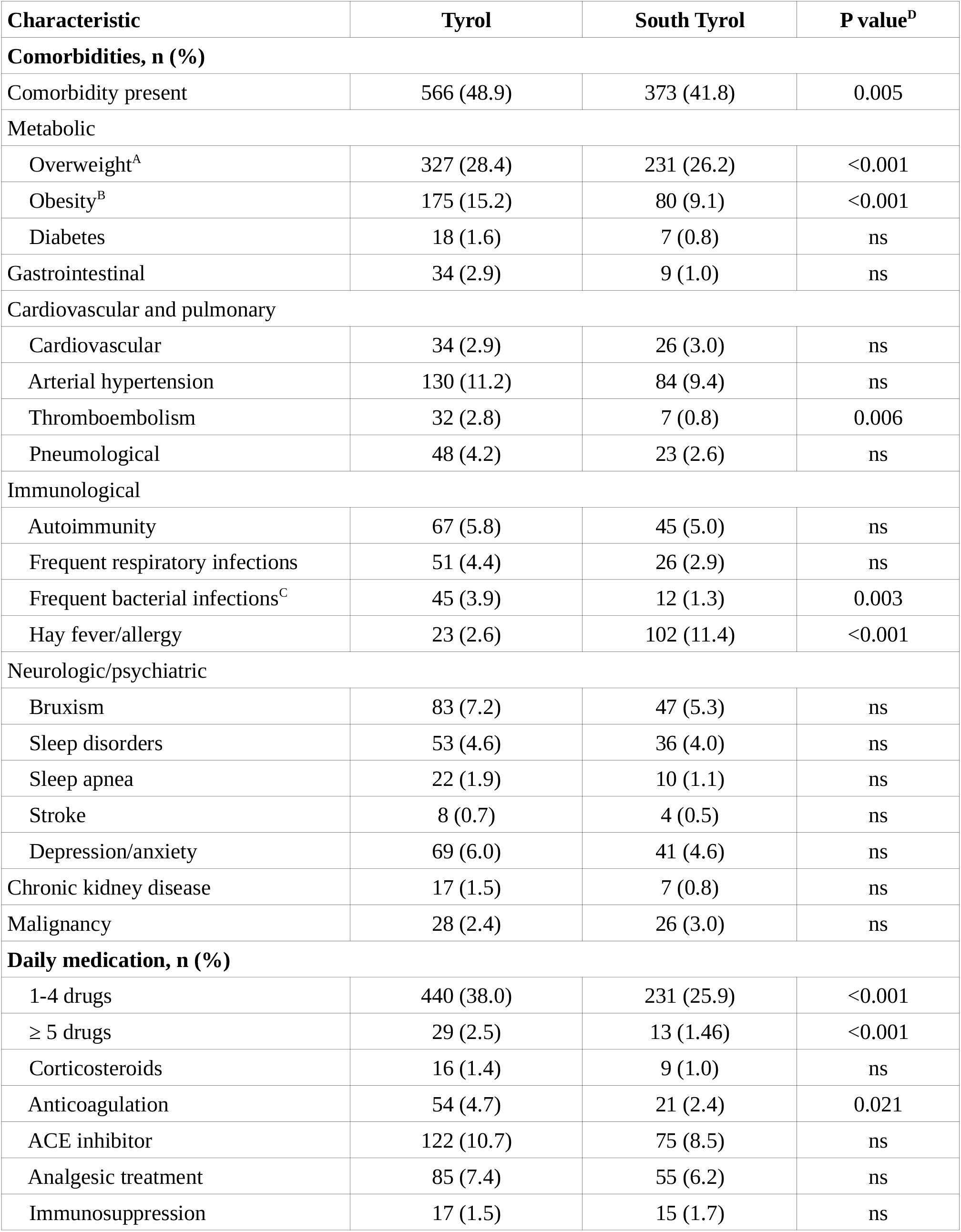
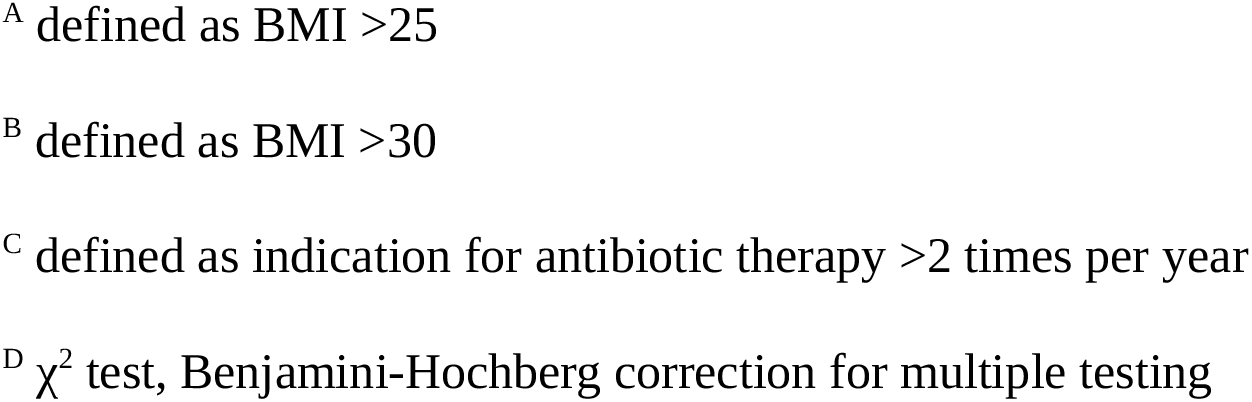
Pre-existing comorbidities and medication in the study cohorts.

| Characteristic | Tyrol | South Tyrol | P value<sup>D</sup> |
| --- | --- | --- | --- |
| **Comorbidities, n (%)** |  |  |  |
| Comorbidity present | 566 (48.9) | 373 (41.8) | 0.005 |
| Metabolic |  |  |  |
| Overweight<sup>A</sup> | 327 (28.4) | 231 (26.2) | <0.001 |
| Obesity<sup>B</sup> | 175 (15.2) | 80 (9.1) | <0.001 |
| Diabetes | 18 (1.6) | 7 (0.8) | ns |
| Gastrointestinal | 34 (2.9) | 9 (1.0) | ns |
| Cardiovascular and pulmonary |  |  |  |
| Cardiovascular | 34 (2.9) | 26 (3.0) | ns |
| Arterial hypertension | 130 (11.2) | 84 (9.4) | ns |
| Thromboembolism | 32 (2.8) | 7 (0.8) | 0.006 |
| Pneumological | 48 (4.2) | 23 (2.6) | ns |
| Immunological |  |  |  |
| Autoimmunity | 67 (5.8) | 45 (5.0) | ns |
| Frequent respiratory infections | 51 (4.4) | 26 (2.9) | ns |
| Frequent bacterial infections<sup>C</sup> | 45 (3.9) | 12 (1.3) | 0.003 |
| Hay fever/allergy | 23 (2.6) | 102 (11.4) | <0.001 |
| Neurologic/psychiatric |  |  |  |
| Bruxism | 83 (7.2) | 47 (5.3) | ns |
| Sleep disorders | 53 (4.6) | 36 (4.0) | ns |
| Sleep apnea | 22 (1.9) | 10 (1.1) | ns |
| Stroke | 8 (0.7) | 4 (0.5) | ns |
| Depression/anxiety | 69 (6.0) | 41 (4.6) | ns |
| Chronic kidney disease | 17 (1.5) | 7 (0.8) | ns |
| Malignancy | 28 (2.4) | 26 (3.0) | ns |
| **Daily medication, n (%)** |  |  |  |
| 1-4 drugs | 440 (38.0) | 231 (25.9) | <0.001 |
| ≥ 5 drugs | 29 (2.5) | 13 (1.46) | <0.001 |
| Corticosteroids | 16 (1.4) | 9 (1.0) | ns |
| Anticoagulation | 54 (4.7) | 21 (2.4) | 0.021 |
| ACE inhibitor | 122 (10.7) | 75 (8.5) | ns |
| Analgesic treatment | 85 (7.4) | 55 (6.2) | ns |
| Immunosuppression | 17 (1.5) | 15 (1.7) | ns |

<sup>A</sup> defined as BMI >25
<sup>B</sup> defined as BMI >30
<sup>C</sup> defined as indication for antibiotic therapy >2 times per year
<sup>D</sup> $\chi^2$ test, Benjamini-Hochberg correction for multiple testing

**Table 3.** Characteristics of acute and post-acute COVID-19 course.

| Characteristic | Tyrol | South Tyrol | P value |
| --- | --- | --- | --- |
| Acute COVID-19 |  |  |  |
| Contact with an infected person, n (%) | 712 (61.7) | 593 (66.6) | 0.03<sup>A</sup> |
| Quarantine duration, n (%) |  |  |  |
| 0-10 days | 536 (47.4) | 52 (5.9) | <0.001<sup>A</sup> |
| 11-14 days | 396 (35.0) | 72 (8.1) |  |
| ≥ 15 days | 198 (17.5) | 762 (86) |  |
| Period of SARS-CoV-2 infection, n (%) |  |  |  |
| Spring 2020 | 309 (26.7) | 144 (16.1) | <0.001<sup>A</sup> |
| Summer/Fall 2020 | 789 (68.2) | 484 (54.2) |  |
| Winter/spring 2020/21 | 59 (5.1) | 265 (29.7) |  |
| Subjective perception of acute COVID-19, n (%) |  |  |  |
| Common cold-like | 289 (25.4) | 248 (28.3) | <0.001<sup>A</sup> |
| Influenza-like | 235 (20.7) | 147 (16.8) |  |
| Gastroenteritis-like | 47 (4.1) | 62 (7.1) |  |
| Not-experienced before | 565 (49.7) | 418 (47.8) |  |
| Weight loss, n (%) | 555 (48) | 355 (39.8) | ns<sup>A</sup> |
| Contact with physician during quarantine, n (%) | 482 (41.8) | 463 (51.8) | <0.001<sup>A</sup> |
| Symptomatic COVID-19 therapy, n (%) |  |  |  |
| None | 864 (74.7) | 593 (66.4) | <0.001<sup>A</sup> |
| Anti-pyretic | 259 (22.4) | 257 (28.8) | 0.003<sup>A</sup> |
| Anbtibiotic | 83 (7.2) | 94 (10.5) | 0.02<sup>A</sup> |
| Acute symptom number, median (IQR) |  |  |  |
| Overall | 13 (9-18) | 13 (7-18) | ns<sup>B</sup> |
| NIP<sup>C</sup> | 8 (6-10) | 8 (6-10) | ns<sup>B</sup> |
| MOP<sup>D</sup> | 3 (1-5) | 3 (1-6) | ns<sup>B</sup> |
| Symptom persistence, n (%) |  |  |  |
| absent | 96 (8.3) | 110 (12.3) | 0.012<sup>A</sup> |
| 1-3 days | 43 (3.7) | 28 (3.1) |  |
| < 1 week | 66 (5.7) | 58 (6.5) |  |
| < 2 weeks | 130 (11.2) | 92 (10.3) |  |
| < 4 weeks | 272 (23.5) | 165 (18.5) |  |
| ≥ 4 weeks | 550 (47.6) | 440 (49.3) |  |
| Post-acute COVID-19 |  |  |  |
| Presence of persistent COVID-19 symptoms, n (%) | 550 (47.6) | 440 (49.3) | ns<sup>A</sup> |
| Symptom relapse, n (%) | 368 (31.9) | 86 (20.9) | <0.001<sup>A</sup> |
| Rehabilitation after COVID-19, n (%) | 14 (1.2) | 7 (0.8) | ns<sup>A</sup> |
| Subjective need for rehabilitation, n (%) | 196 (17.0) | 117 (13.2) | 0.044<sup>A</sup> |
| Subjective complete convalescence, n (%) | 624 (54.0) | 563 (63.3) | <0.001<sup>A</sup> |
| Subjective physical performance loss, n (%) |  |  |  |
| 0-25% | 860 (74.7) | 674 (76.2) | ns<sup>A</sup> |
| 26-50% | 202 (17.5) | 145 (16.4) |  |
| 51-75% | 72 (6.3) | 54 (6.1) |  |
| 76-100% | 17 (1.5) | 11 (1.2) |  |
| Persistent symptom number, median (IQR) |  |  |  |
| Overall | 0 (0-3) | 0 (0-3) | ns<sup>B</sup> |
| HAP<sup>E</sup> | 1 (0-2) | 0 (0-2) | ns<sup>B</sup> |
| FAP<sup>F</sup> | 1 (0-3) | 1 (0-3) | ns<sup>B</sup> |
| MOP<sup>D</sup> | 1 (0-4) | 1 (0-4) | ns<sup>B</sup> |

<sup>A</sup> $\chi^2$ test, Benjamini-Hochberg correction for multiple testing
<sup>B</sup> Mann-Whitney U test, Benjamini-Hochberg correction for multiple testing
<sup>C</sup> Non-specific infection phenotype
<sup>D</sup> Multi-organ phenotype
<sup>E</sup> Hypo-/anosmia phenotype
<sup>F</sup> Fatigue phenotype

### Definitions and variable stratification

Respondents retrospectively assigned their symptoms to pre-defined duration classes (absent, present for 1 – 3 days, ≤ 1 week, ≤ 2 weeks, ≤ 4 weeks, ≤ 3 months, ≤ 6 months and > 6 months). The symptoms were classified as (1) acute, when present in the first two weeks after clinical onset, (2) sub-acute when present 2 – 4 weeks after clinical onset and (3) persistent when lasting ≥ 4 weeks. For modeling, symptom numbers were stratified by quartiles (**Supplementary Table S2**).

Perception of acute infection, symptom relapse, subjective convalescence and rehabilitation need were surveyed as single questions each. Self-reported percent loss of physical performance following COVID-19 was stratified as 0%, ≤ 25%, ≤ 50%, ≤ 75% and > 75%. The detailed variable stratification scheme is shown in the **Supplementary Methods** and **Supplementary Table S1**.

### Statistical analysis

Statistical analysis was performed with R 4.0.5. The analysis pipeline is available at https://github.com/PiotrTymoszuk/health-after-COVID19-analysis-pipeline.

Frequency changes were assessed with χ2 test and differences in medians with Mann-Whitney U or Kruskal-Wallis test, as appropriate. Symptom number kinetics was analyzed with mixed-effect Poisson regression.(12) Symptom phenotypes were defined by PAM (partitioning around medoids) clustering and simple-matching distance.(13, 14) Subsets of long COVID subjects were defined by DBSCAN algorithm and Manhattan distance.(15, 16)

Single-feature (uni-variate) correlation was investigated with gender- and age-weighted Poisson or logistic modeling (see: **Supplementary Table S6** for weighting schemes) with the observation time confounder (diagnosis to survey completion). Pooled Tyrol/South Tyrol regression estimates presented in the text were obtained by the inverse variance method.(17)

Multi-parameter modeling was done with LASSO (least absolute shrinkage and selection operator) 10-fold cross-validated age- and gender-weighted Poisson or logistic regression.(18) LASSO model prediction was evaluated with receiver-operator characteristic (ROC). P values were corrected for multiple testing with Benjamini-Hochberg method.(19) For more details on statistics, see **Supplementary Methods**.

## Results

### Study population

Overall, 2065 individuals in Tyrol and 1075 in South Tyrol participated in the online survey. After exclusion of hospitalized respondents (n = 84 and n = 83, respectively) and questionnaires with an observation period < 28 days (n = 741 and n = 56, respectively), 1157 Tyrol and 893 South Tyrol surveys were eligible for further data analysis (**Figure 1**).

The median COVID-19 diagnosis – survey time was 79 days (IQR: 40 – 175) in Tyrol and 96 days (IQR: 60 – 138) in South Tyrol (p = 1.5×10^−7^). The fractions of respondents with the follow-up time > 180 days ranged between 17.2% (South Tyrol) and 23.1% (Tyrol, p = 3.2×10^−22^) (**Table 1**). Significantly more respondents in South Tyrol (29.7%) than in Tyrol (5.1%, p = 1.9×10^−50^) were diagnosed for SARS-CoV2 infection in 2021 during the outbreaks mainly caused by alpha and beta variants of concern (**Table 3**).(20)

In both collectives, individuals > 65 years were under-represented (5.61 to 5.71%, p = 0.012) with the median participant age of 43 years (IQR: 31 – 53) in Tyrol and 45 years (IQR: 35 – 55) in South Tyrol (**Table 1)**. Females comprised 65.1 -68.3% of the analyzed populations, which was significantly more than in the generalized convalescent population in Tyrol (30 – 54 years, study: 65.7% female, population: 50.6%, p = 9.1×10^−1^) and Italy (30 – 60 years, study: 68.3% female, population: 51.6%, p = 1.0×10^−14^).(21, 22) As expected for the age distribution, employment rate was 81.2 – 81.5% with a substantial over-representation of employees from healthcare (Tyrol: 25.9%, South Tyrol: 20.1%) and educational settings (Tyrol: 12.6%, South Tyrol: 13.3%). Notably, multiple sociodemographic characteristics like employment sectors, education, language structure and residence in the region’s capital area differed significantly between the study collectives (**Table 1**).

41.8 – 48.9% of the study populations reported pre-existing comorbidities and this fraction was significantly lower in the South Tyrol cohort (p = 0.005). The most frequent comorbidities were overweight (BMI 25 – 30 kg/m^2^; 26.2 – 28.4%), hay fever (11.4 – 18%), obesity (BMI > 30 kg/m^2^, 9.08 – 15.2%), arterial hypertension (9.41 – 11.2%), bruxism (5.26 – 7.17%) and depression/anxiety disorders (4.59 – 5.96%) (**Table 2**).

### Characteristics of acute COVID-19 and recovery trajectories

8.3 – 12.3% of participants experienced an asymptomatic SARS-CoV-2 infection, which is below the estimates of the Austrian (16.5% - 26.9%) and Italian (up to 50%) health authority (**Table 3**). (22, 23)

Almost half of the symptomatic participants described acute COVID-19 as a condition ‘not experienced before’ (47.8 – 49.7%), followed by ‘ common cold-like’ (25.4 – 28.3%), ‘influenza (flu)-like’ (16.8 – 20.7%) or ‘gastroenteritis-like’ (4.1 – 7.1%) illness. In most participants (52.8 – 60%), self-reported severe illness perception was limited to one week, for 18 – 27.3% the feeling of severe sickness persisted for longer than 2 weeks, i. e. the duration of the official quarantine. In 29.5 – 32.2% of the convalescents the queried symptoms resolved within two weeks. However, nearly half of the participants (Tyrol: 47.6%, South Tyrol: 49.3%) suffered from at least one persisting symptom for ≥ 28 days. Notably, 20.9 – 31.9% experienced symptom relapse (**Figure 2A, Supplementary Figure S1B, Table 3**).

**Figure 2.**
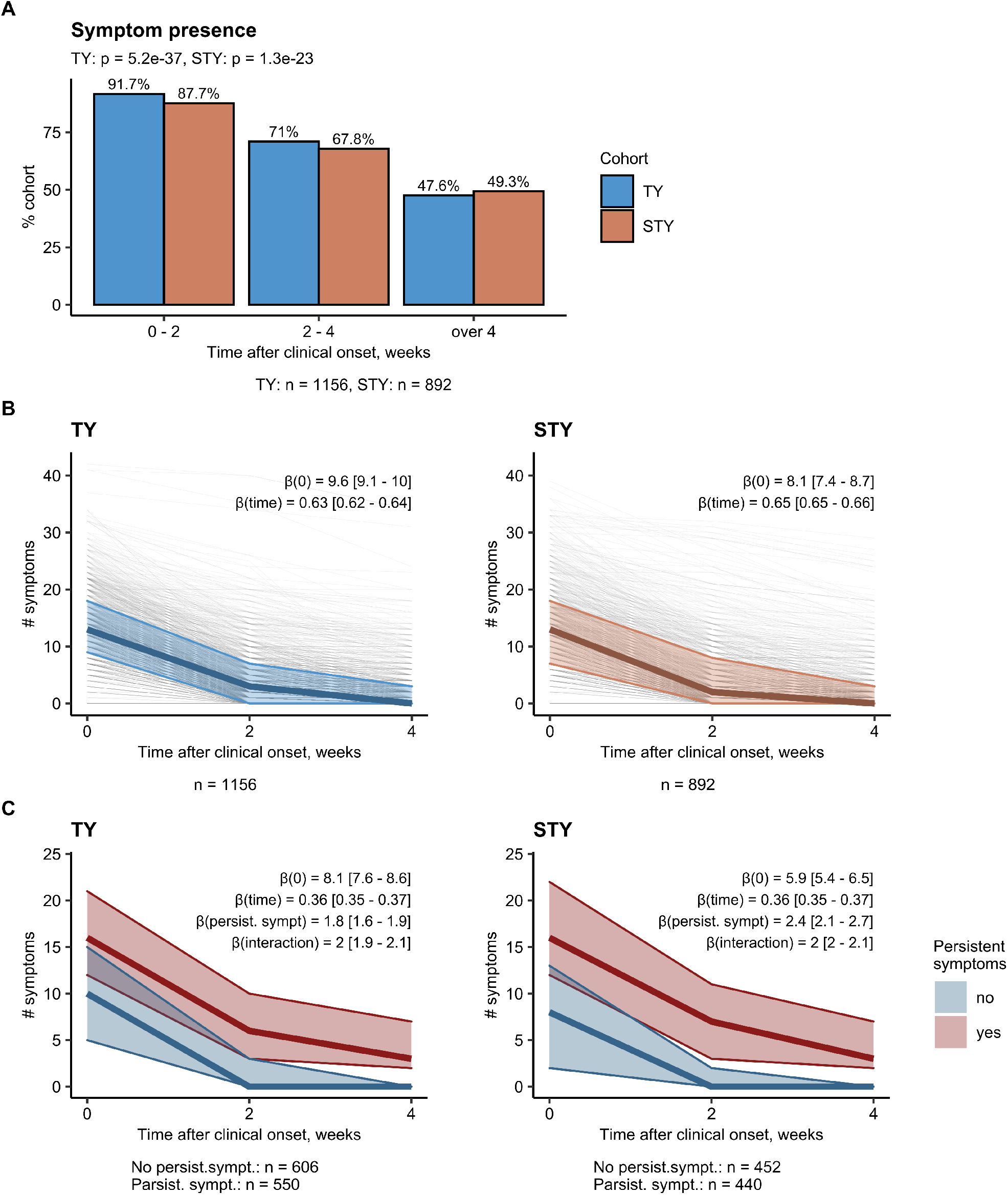
Prevalence and resolution of the symptom number. Individual trajectories of the numbers of disease symptoms (44 symptoms surveyed in total) in the first 2, 2-4 and 4 weeks or longer after symptom onset. **(A)** Percentage of symptomatic participants in time. **(B)** Number of symptoms in the entire study population. **(C)** Number of symptoms in the subsets with and without persistent symptoms (4 weeks or longer after symptom onset). Thin grey lines: individual symptom number trajectories, thick color line: median symptom count, color ribbon: IQR. Statistical significance was determined by mixed-effect Poisson modeling. Model estimates with 95% CI and p values are indicated in the plot, number of complete responses is shown under the plots. TY: Tyrol, STY: South Tyrol cohort.

The number of acute COVID-19 symptoms present in the first two weeks (Tyrol, median: 13 (IQR: 9 – 18), South Tyrol: 13 (IQR: 7 – 18) out of 44 symptoms queried) and the weekly symptom count resolution rate of 35% were comparable in the study collectives (**Figure 2B, Table 3**). Interestingly, study participants suffering from at least one persistent symptom had on average both, an acute symptom burden twice as high (β_persist.sympt._ = 1.8 [95%CI: 1.6 – 1.8] to 2.4 [95%CI: 2.1 – 2.7]) and a two times slower resolution rate (β_interaction_ = 2 [95%CI: 1.9 – 2.1] to 2 [95%CI: 2 – 2.1]) (**Figure 2C**).

### Prevalence of acute and persistent COVID-19 symptoms

Besides highly frequent non-specific infection symptoms (fatigue, headache, joint pain, myalgia, diminished appetite, fever), manifestations of respiratory dysfunction: tachypnoea (51 – 57%), chest pain (41 – 47%) and dyspnoea (28 – 34%) were present in a considerably large percentage of the symptomatic outpatients during acute SARS-CoV2 infection (**Figure 3, Supplementary Figure S1 – S2**).

**Figure 3.**
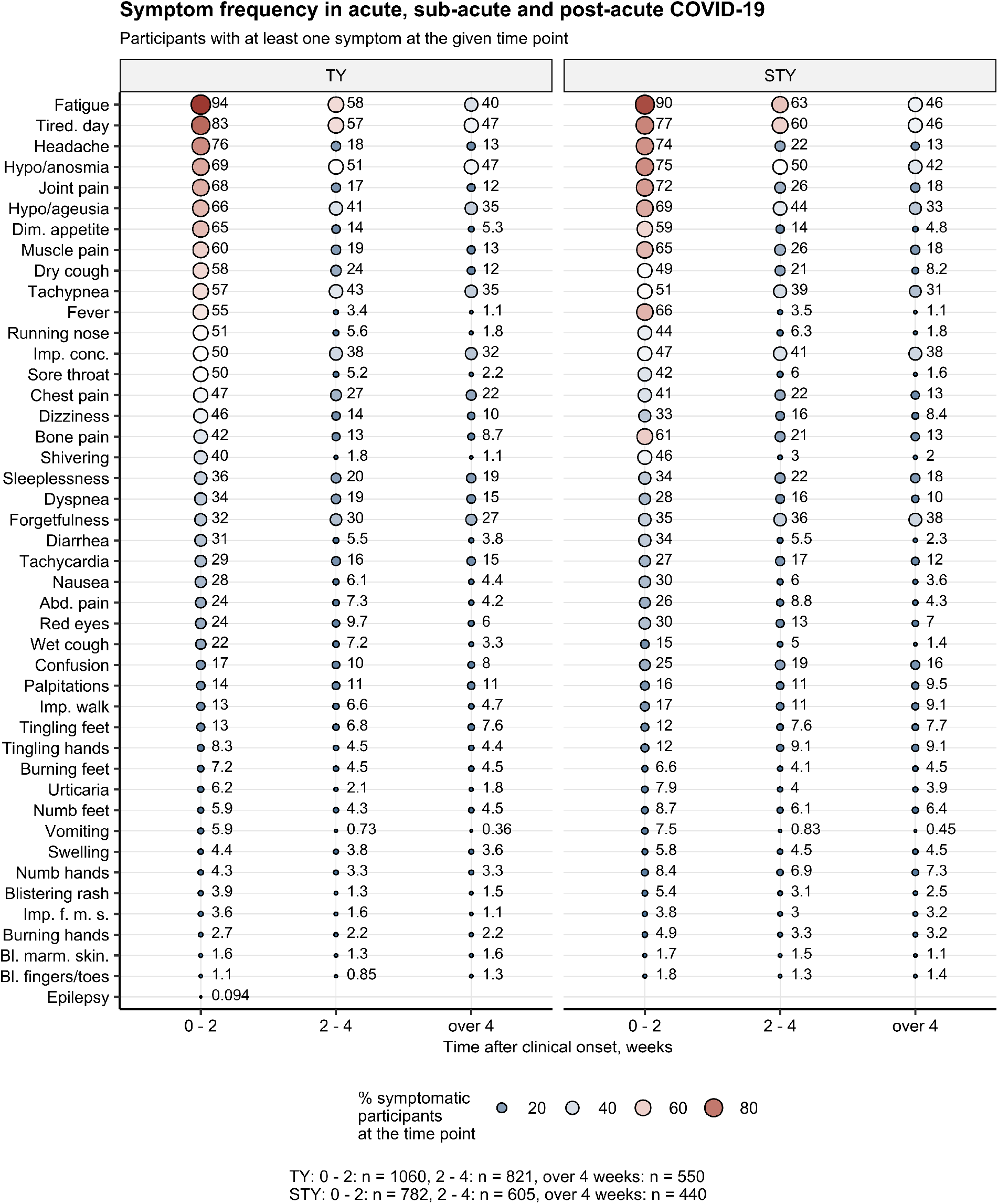
Symptom frequency in acute, sub-acute and post-acute phase of COVID-19. Symptom frequencies were calculated as percentages of the individuals reporting at least one symptom in the first two, two to four weeks and four weeks or longer after symptom onset. % within the symptomatic subset are presented as a bubble plot. Point size and color represents the percentage. Numbers of complete observations are indicated below the plot. tired. day: tiredness at day, imp. conc.: impaired concentration, abd. pain: abdominal pain, imp. walk: impaired walk, dim. appetite: diminished appetite, imp.f.m.s: impaired fine motor skills, TY: Tyrol, STY: South Tyrol cohort.

Frequency of nearly all symptoms and especially cold- or flu-like complaints declined significantly over time (**Figure 3, Supplementary Figure S1, Supplementary Table S3**). In contrast, resolution of fatigue (40 – 46% of subjects with persistent symptoms), daytime tiredness (46 – 57%), hyposmia/anosmia (42 – 47%), taste disorder (33 – 35%) and tachypnoea (31 – 35%) were substantially delayed. Those features together with concentration (32 – 38%) and memory deficits (27 – 38%) represented the predominant manifestations of post-acute sequelae (**Figure 3, Supplementary Figures S1 – S2**).

Furthermore, 13.7 – 14% of participants reported loss of hair and 4.38 – 5.21% a weight reduction > 5 kg during convalescence. 23.8 – 25.3% of respondents reported a physical performance loss exceeding 25% and for 7.3 – 7.7% it was even more than 50%. Over one third of the study populations (36.7 – 46%) reported on an incomplete recovery and 13.2 – 17% declared a need for rehabilitation (**Table 3**).

### Patterns of acute and persistent COVID19 symptoms

Applying cluster analysis, two reproducible acute COVID-19 symptom patterns, further termed ‘phenotypes’, were identified in both cohorts.(13) The, non-specific infection phenotype‘ (NIP) consisted of symptoms characteristic for upper respiratory tract infections such as rhinitis, sore throat, dry cough and fatigue, along with smell and taste disorders. The ‘multi-organ phenotype’ (MOP) included closely co-occurring symptoms of the lower respiratory tract and wide-ranged neurological, gastrointestinal, cardiovascular and dermatological manifestations (**Figure 4, Supplementary Figure S3, Supplementary Table S4**). Of note, neither the NIP nor MOP symptom count differed between the study cohorts (**Table 3**).

**Figure 4.**
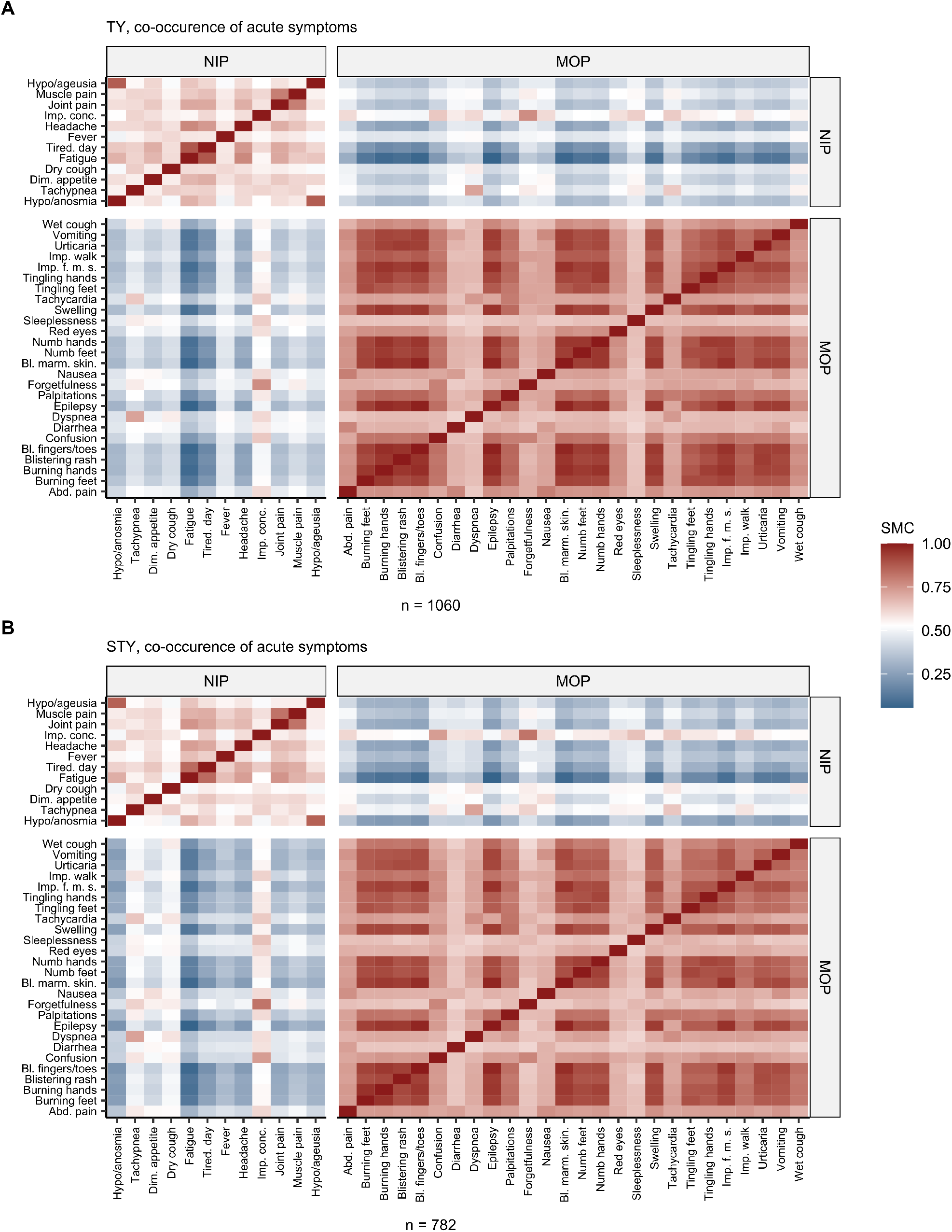
Co-occurrence and clustering of acute COVID-19 symptoms. Simple matching distances between the symptoms of COVID-19 present in the first two weeks after symptom onset were subjected to clustering with PAM (partitioning around medoids) algorithm. Consensus clusters of the same symptoms in both cohorts were termed phenotypes. Values of simple matching coefficient between each two symptoms assigned to the consensus non-specific infection phenotype (NIP) and multi-organ phenotype (MOP) were presented as a heat map. Numbers of complete observations are indicated below the plots. Tired. day: tiredness at day, imp. conc.: impaired concentration, abd. pain: abdominal pain, imp. walk: impaired walk, dim. appetite = diminished appetite, imp.f.m.s: impaired fine motor skills.

By an analogical procedure, we found three phenotypes of persistent COVID-19 manifestations: (1) ‘hyposmia/anosmia phenotype’ (HAP) encompassing closely co-occurring smell and taste disorder, (2) ‘fatigue phenotype’ (FAP) including fatigue, tiredness, memory and concentration deficits, and (3) ‘multi-organ phenotype’ (MOP) comprising a combination of pulmonary, gastrointestinal, neuro-cognitive, psychosomatic and cardiovascular disorders **(Figure 5, Supplementary Table S4)**.

**Figure 5.**
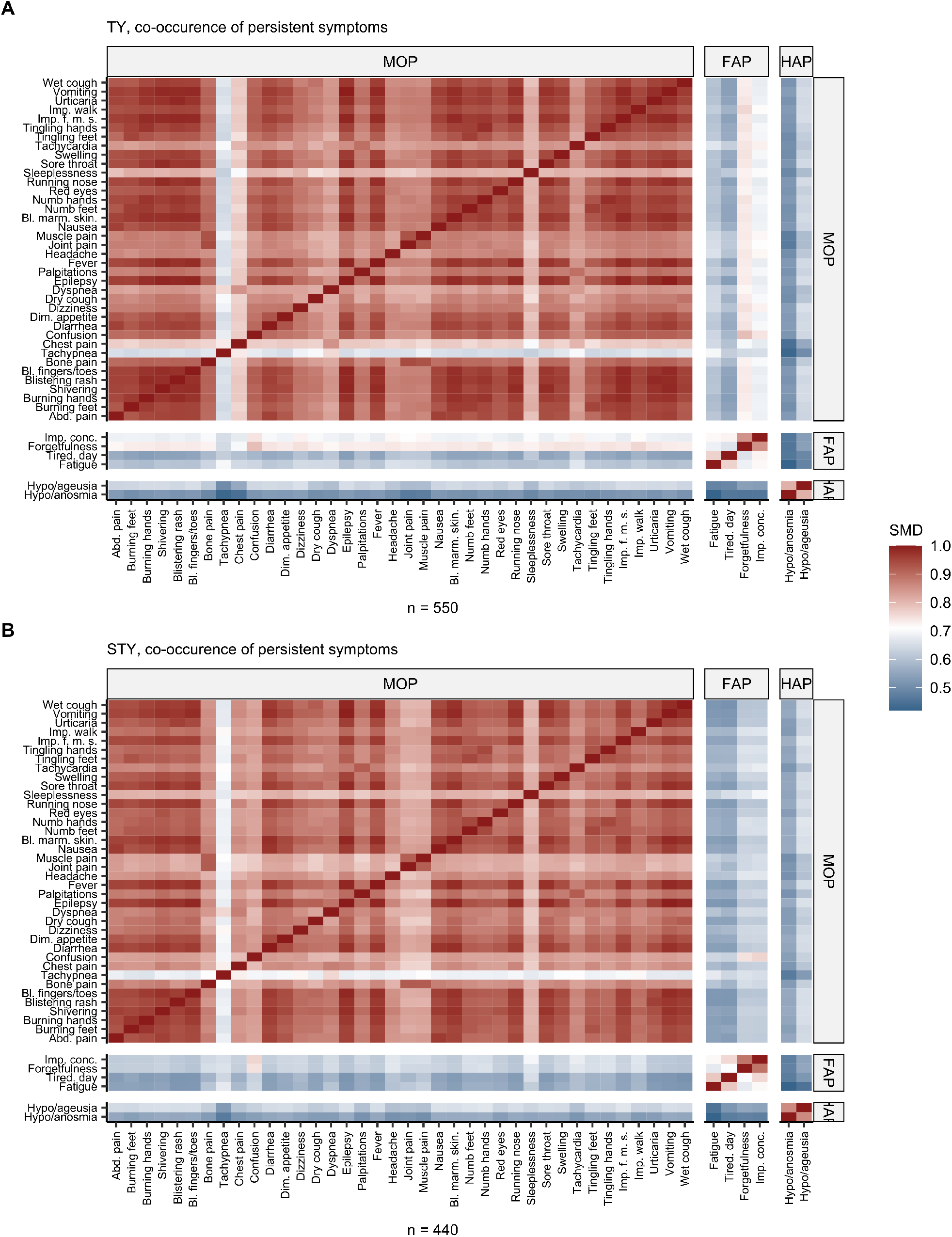
Co-occurrence and clustering of persistent COVID-19 symptoms. Simple matching distances between the symptoms of COVID-19 present for four weeks or longer after symptom onset were subjected to clustering with PAM (partitioning around medoids) algorithm. Consensus Clusters of the same symptoms in both cohorts were termed phenotypes. Values of simple matching coefficient between each two symptoms assigned to the consensus hyposmia/anosmia phenotype (HAP), fatigue phenotype (FAP) and multi-organ phenotype (MOP) were presented as a heat map. Numbers of complete observations are indicated below the plots. Tired. day: tiredness at day, imp. conc.: impaired concentration, abd. pain: abdominal pain, imp. walk: impaired walk, dim. appetite: diminished appetite, imp.f.m.s: impaired fine motor skills.

Based on the number of persistent HAP, FAP and MOP features, three distinct subsets of the individuals suffering from persistent COVID-19 symptoms were characterized, which differed primarily by the count of the HAP taste and smell disorders.(15) Conspicuously, the subset suffering from one HAP complaint (13.2 – 18.5%) displayed the lowest FAP and MOP persistent symptom burden. In contrast, the largest HAP-negative subset (30.9 – 48.7%) had the highest burden of multi-organ and fatigue phenotype complaints (**Supplementary Figure S5 – S6**).

### Factors linked to the severity of acute COVID-19

By means of univariate, age-, sex and observation time-adjusted serial modeling we found 15 factors significantly correlating with the acute symptom count in both study collectives (**Supplementary Figure S7A**). Among them, hallmarks of subjective infection severity: contact with a physician, need of symptomatic treatment with anti-pyretic or antibiotic drugs and duration of the home isolation > 14 days were associated with a 30 – 46% increase of the symptom number in the study populations. Additionally, measures of overall health status before infection such as ≥3 comorbidities, obesity, pulmonary disease and > 2 respiratory infections per year, depression or anxiety and sleep disorders were linked to > 10% higher acute COVID-19 symptom burden. In turn, a 23 – 29% lower symptom count was observed in males and in participants not having received any symptomatic therapy during acute SARS-CoV2 infection (**Supplementary Figure S7B, Supplementary Tables S5 – S7**, for pooled β estimates, see: **Supplementary Table S8**). By multi-parameter LASSO modeling, the readouts for subjective severe illness perception factors linked to a higher acute symptom burden were: need to consult a physician (β = 1.18 – 1.24), anti-pyretic treatment (1.02 – 1.06) and ≥ 3 pre-existing co-morbidities (1.05 – 1.12). Opposite, no symptomatic therapy (β = 0.93 – 0.96) and male sex (0.89 – 0.93) could be corroborated as independent co-variates linked to low risk for perception of severe illness (**Figure 6, Table S8**).(18)

**Figure 6.**
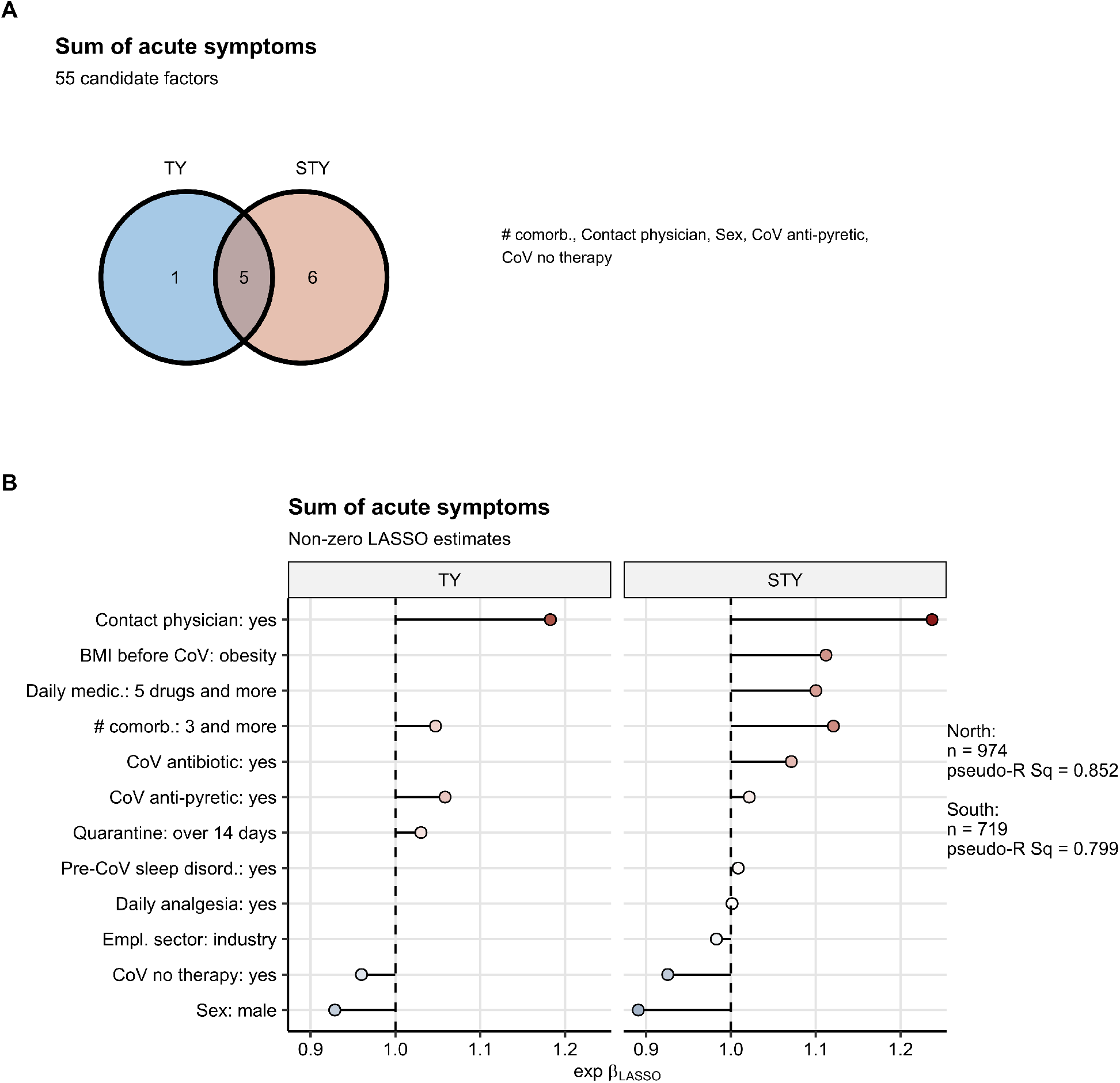
Identification of independent factors associated with the symptom burden of acute COVID-19. Correlation of 55 candidate factors with the sum of acute COVID-19 symptoms (first two weeks after symptom onset) was investigated by sex- and age-weighted LASSO (least absolute shrinkage and selection operator) Poisson regression in the Tyrol and South Tyrol cohorts. **(A)** Numbers of significant independent factors associated with the acute symptom burden in each of the study populations presented in a Venn plot. Common significant factors are listed next to the plot. **(B)** Values of non-zero regression coefficients of the significant factors correlating with the acute symptom burden presented in the plot. Numbers of complete observations and pseudo-R^2^ statistics for each LASSO model are presented next to the plot. CoV antibiotic/CoV anti-pyretic/CoV no therapy: antibiotic/anti-pyretic/no symptom-specific treatment during acute COVID-19, # comorb.: number of pre-existing co-morbidities, #CoV in household: number of COVID-19 cases in the household, Daily medic.: daily medication, Pre-CoV sleep disord.: sleep disorders before COVID-19, Freq. resp. inf.: frequent respiratory infections, Empl. sector: employment sector, Inf. contact: contact with an infected person before COVID-19, Auth. support: support from authorities during quarantine, Contact physician: contact with a physician during quarantine, Empl. status: employment status.

### Factors linked to symptom persistence or relapse and major physical loss of performance

By uni-variate modeling, 30 factors correlating with the risk of developing persistent symptoms, relapse and more than 50% physical performance loss were identified (**Supplementary Figure S8**).

Especially, the overall number of symptoms, count of MOP manifestations and fatigue during acute SARS-CoV2 infection were associated with > 6-fold risk of symptom persistence, relapse and physical performance loss following COVID-19. Neuro-cognitive symptoms present in acute COVID-19 such as forgetfulness, concentration deficits, confusion and dizziness constituted another group of shared unfavorable risk factors linked to linked to an > 3-fold risk of the investigated long-COVID features. Furthermore, acute cardiopulmonary manifestations such as tachycardia, heart palpitations, shortness of breath and dyspnea were associated with an > 2.5-fold increase in risk of developing the investigated long COVID-19 features. Interestingly, male sex and no symptom-specific therapy during acute SARS-CoV2 infection were associated with a 34 – 45% reduced risk of development of persistent symptoms and relapse but not with significantly physical impairment (**Supplementary Figures S9 – S11, Table S7**, for pooled β estimates, see: **Supplementary Table S9**).

In multi-parameter LASSO modeling, acute forgetfulness (OR = 1.52 – 1.88), dizziness (1.01 – 1.09) and tachypnea (1.38 – 1.5) were corroborated as independent unfavorable risk factors of symptom persistence in both study cohorts together with acute loss of smell (1.3 – 1.5), taste disorders (1.06 – 1.31) and loss of hair (1.07 – 1.08). Additionally, > 180 day time interval between COVID-19 diagnosis and survey completion was also associated with the higher symptom persistence risk (1.02 – 1.41). Interestingly, in the Tyrol cohort, daily ACE inhibitor intake was linked to a significantly diminished symptom persistence risk (uni-variate, Tyrol: OR = 0.59 [95%CI: 0.38 – 0.904], LASSO: OR = 0.833) (**Figure 7, Supplementary Table S10**). Applying the LASSO approach to the relapse and major physical impairment risk, the overall number of acute symptoms (> 75^th^ percentile, relapse: OR = 1.14 – 1.56, performance loss: 1.23 – 1.26) and chest pain during acute infection (relapse: OR = 1.14 – 1.56, OR = 1.02) were discerned as independent unfavorable risk-modifying features both in the Tyrol and South Tyrol cohorts. Additionally, the number of acute multi-organ symptoms (> 75^th^ percentile: OR = 1.31 – 1.38) proved a shared independent unfavorable factor specifically for the performance impairment risk (**Supplementary Figures S12 – S13, Supplementary Table S10**).

**Figure 7.**
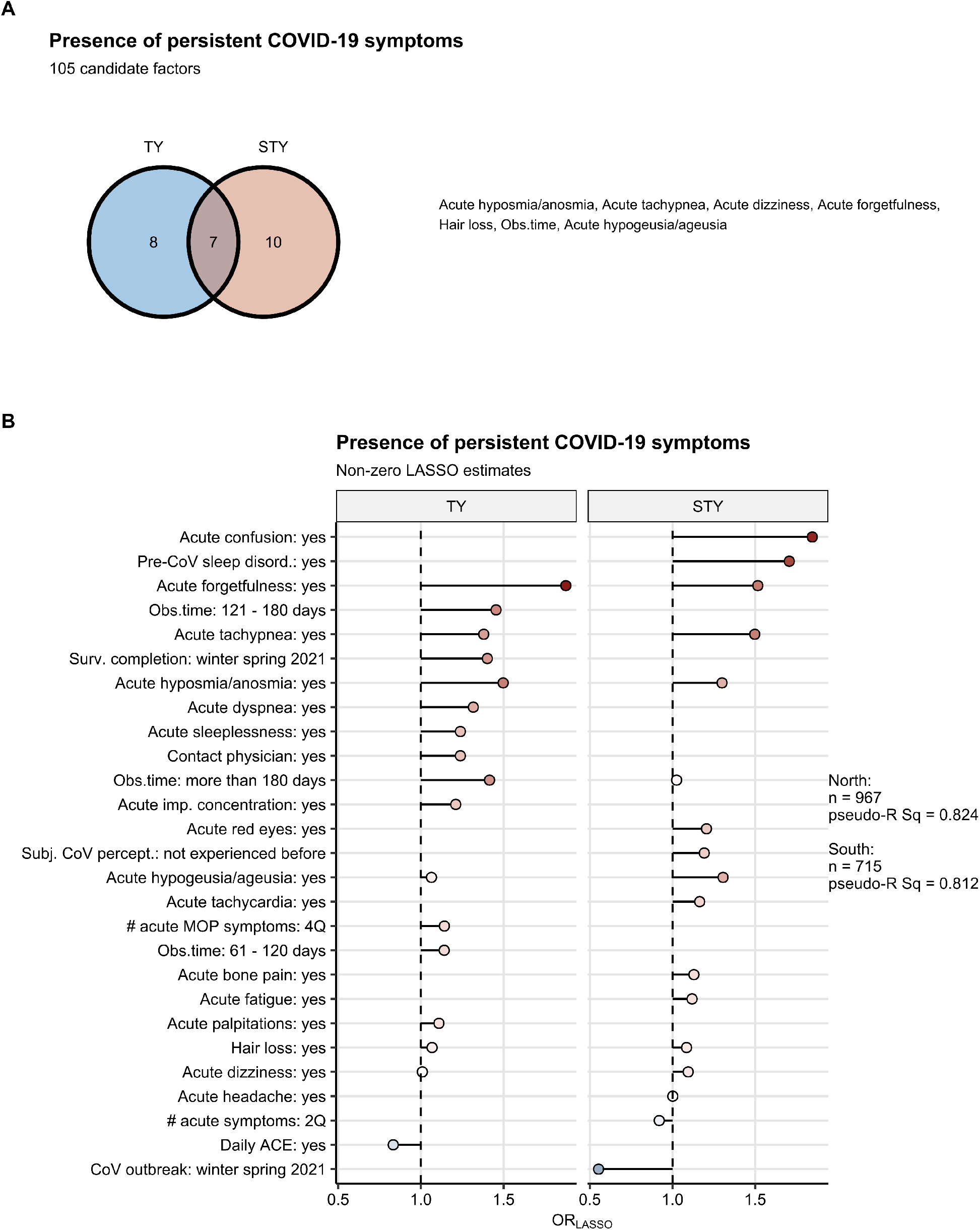
Identification of independent factors associated with the presence of persistent COVID-19 symptoms. Correlation of 105 candidate factors with the risk of developing at least one persistent symptom (four weeks or longer after symptom onset) was investigated by sex- and age-weighted LASSO (least absolute shrinkage and selection operator) logistic regression in the North and South Tyrol cohorts. **(A)** Numbers of significant independent factors associated with the risk of developing persistent symptoms in each of the study populations presented in a Venn plot. Common significant factors are listed next to the plot. **(B)** Values of non-zero regression coefficients of the significant factors correlating with the risk of developing persistent symptoms presented in the plot. Numbers of complete observations and pseudo-R^2^ statistics for each LASSO model are presented next to the plot. Obs.time: observation time (test to participation), Pre-CoV sleep disord.: sleep disorders before COVID-19, Surv. completion: survey completion, Acute imp. concentration: acute impaired concentration, Subj. CoV percept.: subjective perception of acute COVID-19. # acute MOP symptoms: sum of acute multi-organ phenotype symptoms, # acute symptoms: sum of acute COVID-19 symptoms, Daily ACE: daily intake of ACE inhibitors, CoV outbreak: SARS-CoV2 outbreak, Acute bl. marm. skin.: acute blue marmorate skin, 2Q: second quartile, 4Q fourth quartile.

Finally, the LASSO models developed in the Tyrol cohort (**Supplementary Table S11**) enabled reliable predictions of the persistence and relapse of symptoms and major physical impairment in the South Tyrol collective (**Supplementary Figure S14**). This indicates that a similar set of clinical and demographic features determines the propensity of long COVID in both study collectives.

## Discussion

This international survey encompassed 2050 participants in two cohorts differing in recruitment mode, ethnics, education and employment structure, observation time, co-morbidity and, most importantly, in the health care systems and pandemic management. Hence, the results enabled us to delineate features of acute COVID-19 and the process of recovery in a non-hospitalized population constituting the great majority of SARS-CoV2 cases.

Regarding the acute presentation of COVID-19 the study collectives displayed a spectrum of symptoms with fatigue, headache, hyposmia/anosmia/dysgeusia, joint pain, dry cough, myalgia, rhinitis, fever and diarrhea as leading complaints. Interestingly, multi-morbidity, in particular obesity, lung disease, frequent respiratory infections, sleep or depressive/anxiety disorder as well as female sex correlated with an increased symptom number as a surrogate of acute COVID-19 severity. The acute manifestations were classified as two distinct co-occuring patterns reproducible in both study collectives. The ‘non-specific infection phenotype’ (NIP) included the most frequent acute symptoms shared by multiple upper-respiratory infections. In turn, the ‘multi-organ phenotype’ (MOP) comprised of wide-ranged neurological, gastrointestinal, cardiovascular and dermatological manifestations, which pertain to acute ‘atypical’ and multi-systemic complaints reported in large-scale studies.(7, 8, 24) Most probably, high MOP density reflects more severe infection and may correlate with the need of professional medical support, as suggested for the patient cluster with frequent abdominal pain and confusion classified here as MOP symptoms.(24) The MOP phenotype was not only detectable in acute SARS-Cov2-infection but also in patients with post-acute symptoms. Mechanistically, acute MOP symptoms density and persistence may pathophysiologically involve viral pneumonia and encephalopathy, hyper-inflammatory immune response and/or pathological coagulation.(25) Thus, further research on the biological mechanisms of long COVID and its heterogeneity is urgently needed.

Clinically, 50% of participants suffered from complaints persisting ≥ 28 days and, hence, fulfilled the definition of ‘long-COVID’.(3) This frequency is located within the reported range (13-76%) of long COVID depending on sample size, observation time and research design.(7, 8, 25, 26) In line with recent evidence, those long COVID subjects had both a two-fold elevated number of acute symptoms and a halved resolution pace as compared to the subgroup without persistent symptoms. (8, 10) Specifically, the resolution of fatigue, tiredness, smell and taste disorders, tachypnea and MOP manifestations such as forgetfulness, tachycardia and confusion were delayed and represented a congruent signature of post-acute sequelae as previously described by others.(5, 8, 10, 25) Furthermore, the overall number of acute infection symptoms, fatigue, smell and taste disorders, acute MOP symptom count and particular MOP features including neuro-cognitive deficits, sleeplessness and cardiopulmonary abnormalities could be proposed as strong correlates of elevated symptom persistence, relapse and major physical impairment risk. Analogically to the previous reports we found lower symptom persistence and recurrence rates in males and subjects having experienced mild acute infection not requiring any symptomatic treatment in univariate modeling approach.(5, 8) However, neither male sex nor mild-course acute COVID not requiring a symptomatic treatment proved independent favorable in the multi-parameter analysis. This suggests a lower frequency of other risk factors in males and association of the mild-course acute COVID-19 with lower overall and MOP symptom burden.

Among long COVID participants, we unravelled three subsets differing in quality and quantity of post-acute manifestations. The minor subset characterized by a single isolated smell or taste disorder demonstrated the lowest count of persistent symptoms and, in particular, hardly any MOP complaints. Conversely, the hypo/anosmia- and hypo/ageusia-free subset was the most affected by persistent symptoms. Whether this observation may be biased by inter-individual perception of smell and taste, as recently described(27), or reflects a yet uncharacterized pathological phenomenon needs to be clarified.

Our study bears limitations. As indicated by the prevalence of long COVID, fraction of females, middle-aged and over-representation of health care workers in the study collectives, the survey might have targeted primarily individuals more affected by the disease and health-aware people of the general convalescent populations. Additionally, the retrospective and cross sectional study character precluded detailed tracking of particular symptom kinetic and relapses. Furthermore, it may explain the significant effect of the observation time on the readouts of long COVID in the multi-parameter modeling. Even though our data are highly consistent for both study populations differing in multiple demographic, socioeconomic and clinical features and the confounding impact of sex, age and observation time was included in the risk modeling, our findings will require an independent prospective validation.

## Conclusion

Herein, we present a comprehensive description of acute and post-acute COVID-19 manifestations in outpatients, the symptom co-occurrence and patient’s heterogeneity in two independent study cohorts. Density and quality of MOP symptoms are tightly associated with the features of long COVID such as symptom persistence, relapse and physical impairment. These findings may help to develop predictive tools to identify individuals at risk for long COVID and establish concepts of therapy and early rehabilitation.

## Data Availability

Fully anonymised participant data and the study protocol are available at request to. The data analysis pipeline is available at: https://github.com/PiotrTymoszuk/health-after-COVID19-analysis-pipeline.

## Author Contribution

GW, RH, RBW, HB, BSU, GN, SS, VR, AP, MA, KC, BB, and JLR designed the study. SS, PT, PW, DA, GR, BH and GP collected the data. PT and DA performed data analysis. PT, SS, DA, AP, VR, KH, KK, GW, RBW, HB, AB, TS, SK, CW, BSU, GP, AH, RH, IT and JLR interpreted the data. SS, PT, DA, VR, AP, IT, GW, RH and JLR wrote the manuscript. All authors critically reviewed the final version of the manuscript.

## Acknowledgments

We acknowledge all the participants of the study for helping to improve our knowledge of health after COVID-19.

## Funding

The study was funded by the Research Fund of the State of Tyrol, Austria (Project GZ 71934, JLR).

## Conflict of interest

The authors declare no conflict of interest related to this study.

## Notes

### Competing Interest Statement

The authors have declared no competing interest.

### Clinical Trial

NCT04661462

### Author Declarations

This study was approved by the institutional review boards of the Medical University of Innsbruck (Austria) (approval number: 1257/2020) and the South Tyrol Province (Italy) (0150701).

