## Supplementary Material for "Phenotyping of acute and persistent COVID-19 features in the outpatient setting: exploratory analysis of an international cross-sectional online survey"

### COVID-19 phenotyping to predict the risk for long COVID in the outpatient setting: results from a cross-sectional online survey

Supplementary Material

Health after COVID-19 in Tyrol study team

2021-08-05

**Data availability** **5**

**Supplementary Tables** **6**

**Supplementary Figures** **19**

**References** **44**

### Supplementary Methods

#### Study design and participants

The study 'Health after COVID-19 in Tyrol' (ClinicalTrials.gov: NCT04661462) was conducted as an international cooperation project between two regions: Tyrol, the first major COVID-19 hotspot in Austria, and the Italian province South Tyrol. Between 30$^{th}$ September 2020 and 5$^{th}$ July 2021, COVID-19 convalescents recovering from SARS-CoV2 infection confirmed by nasal or oral swab PCR or blood antibody test were invited to participate in an anonymized, cross-sectional web-based survey[1] via public media call or contact with a physician. Residence in the study regions and age $\geq$ 16 (Tyrol) or $\geq$ 18 years (South Tyrol) were additional study inclusion criteria.

Analysis exclusion criteria in this report were hospitalization because of SARS-CoV2 infection and the survey completion at less than 28 days after SARS-CoV-2 infection was diagnosed. The scheme of participant inclusion in the study analysis is provided in **Figure 1**. Socioeconomic and demographic baseline characteristic of the study populations are presented in **Table 1**.

This study was approved by the institutional review boards of the Medical University of Innsbruck (approval number: 1257/2020) and of the South Tyrol Province (approval number: 0150701). Digital informed consent was obtained from each participant at the survey start.

#### Measures

The study questionnaire was developed by a multidisciplinary team (infectious disease specialists, pneumologists, internists, neurologists, psychiatrists, dermatologists, general practitioners, public health and rehabilitation physicians). The survey recorded information on demographics (age, sex, height, weight before infection), socioeconomic status (education, profession, employment status, residence, household size), pre-existing comorbidities (25 items), smoking history, daily medication (quantity, major drug types), vaccination status (flu and pneumococci), course of acute SARS-CoV infection (contact with an infected individual, incubation time, quarantine duration, contact with authorities and physicians), presence and duration of COVID-19-associated symptoms (44 items), illness perception, symptom relapse as well as psychosocial health and physical constitution of during COVID-19 convalescence. Study variables with their descriptions are listed in **Supplementary Table S1**. Baseline demographic, socioeconimic and clinical characteristic of the study populations is presented in **Table 1**. Features of acute COVID-19 course in both study cohorts are presented in **Table 2**. Post-acute characteristic of the study populations is shown in **Table 3**.

#### Definitions and variable stratification

Respondents were asked to retrospectively assign the presence and duration classes of acute symptoms (absent, present for 1 - 3 days, up to 1 week, up to 2 weeks, up to 4 weeks, up to 3 months, up to 6 months and > 6 months). The surveyed symptoms were classified as (1) acute, when present in the first two weeks after clinical onset, (2) sub-acute when present two to four weeks after clinical onset and (3) persistent when lasting for four weeks or longer. Based on the self-reported symptom duration data, we calculated individual time intervals until symptom recovery.

Symptom relapse after the initial resolution, subjective convalescence and need for rehabilitation were surveyed with as a one question item each. Illness perception was queried as 'cold-like', 'flu-like', 'gastroenteritis-like' or 'not experienced before/other'. Assessment of physical performance impairment at the time of study completion compared to the time before COVID-19 was based on participants´ subjective estimate stratified as no impairment (0%), $\leq$ 25%, $\leq$ 50%, $\leq$ 75% and > 75% of performance loss.

For modeling tasks, the count of all acute symptoms as well as acute symptoms assigned to the particular phenotypes was stratified by quartiles as presented in **Supplementary Table S2**. Participant's age was stratified with the 30 and 65 y cut-points (young: 0 - 30, middle aged: 31 - 65, elderly: 66 years and more). For the detailed variable stratification scheme, refer to **Supplementary Table S1_**.

#### Statistical analysis

##### Data transformation and visualization

Self-reported demographic, biometric, symptom and follow-up data were analyzed and visualized with R programming suite (version 4.0.3) and *tidyverse* environment.[2] Visualization of the data, modeling and clustering results was done with packages *ggplot2*,[3] *cowplot*,[4] *ggvenn* and *plotROC*.[5] The entire R analysis pipeline is available at https://github.com/PiotrTymoszuk/health-after-COVID19-analysis-pipeline.

##### Descriptive statistic and hypothesis testing

For assessing statistical significance for changes in variable strata frequency or symptom prevalence in time, $\chi^2$ test and $\chi^2$ test for trend were applied. To compare differences in medians of numeric variables, Mann-Whitney U test and Kruskal-Wallis test were used, as appropriate for the group number. The R functions used for descriptive statistic and hypothesis testing are available at https://github.com/PiotrTymoszuk/analysis_tools. If not indicated otherwise, p values were corrected for multiple testing with Benjamini-Hochberg method.[6]

##### Symptom kinetics modeling

In symptom burden modeling, only the participants with the complete set of symptom answers were included. To model kinetics of individual symptom count contraction as a function of time after clinical onset in the entire study cohorts mixed-effect generalized linear modeling (GLM, fixed-effect: numeric time after clinical onset, random effect: participant, assumed residual distribution: Poisson, log link function) was applied with function *glmer()* from *lme4* package.[7]

To compare symptom count resolution rates between the individuals suffering from persistent symptoms and the participants with complete symptom contraction till week four after clinical onset, analogical mixed-effect GLM approach was utilized (GLM, fixed-effects: numeric time after clinical onset, persistent symptom presence and the time:persistent symptom presence interaction, random effect: participant, assumed residual distribution: Poisson, log link function).[7] The value of exponentiated $\beta_{time}$ coefficient was interpreted as an estimate symptom resolution rate, the value of exponentiated $\beta interaction$ was assumed an estimate of symptom contraction rate between the participants with and without persistent symptoms. Estimate significance was assessed with two-sided T test and degrees of freedom calculated with Satterthwaite method.

##### Co-occurrence of symptoms and identification of disease phenotypes

To investigate co-occurrence of acute and persistent symptoms of COVID-19 pairwise simple matching distances (SMD, function *smc()*, package *scrime*)[8] between the symptoms were subjected to clustering with PAM (partitioning around medoids, function *pam()*, package *cluster*) algorithm.[9] In each clustering task, only the participants with COVID-19 symptoms at the given time point (i.e. first two or four weeks or longer) were included. The decision on the optimal cluster number in each cohort was based on 'elbow' of the sums-of-square curve (function *fviz_nbclust()*, package *factoextra*, **Supplementary Figure S3A** and **Supplementary Figure S4A**). The R tools for clustering analysis are available at https://github.com/PiotrTymoszuk/cluster_tools.

As presented in **Supplementary Figure S3B** and **Supplementary Figure S4B**, although the clustering structure was very similar in both study populations, the symptom cluster members differed slightly. To account for that, acute and persistent COVID-19 phenotypes were defined as the consensus set of symptoms assigned to the same cluster in both study cohorts. By this means, two phenotypes of acute (NIP: non-specific infection phenotype and MOP: multi-organ phanotype) and three phenotypes of persistent (HAP: hyposmia/anosmia phenotype, FAP: fatigue phenotype, MOP: multi-organ phenotype) COVID-19 symptoms were defined.

#### Subsets of subjects suffering from persistent symptoms

To identify diverse subsets of the study participants suffering from persistent COVID-19 symptoms, Manhattan distances (function *distance()*, package *philentropy*)[10] between the affected individuals in respect to normalized sums of HAP (hyposmia/anosmia phenotype), FAP (fatigue phenotype) and MOP (multi-organ phenotype) symptoms were subjected to DBSCAN clustering (function *dbscan()*, package *dbscan*).[11] The minPts argument was set to five based on the $> 2^{data\ dimension}$ rule. The decision on the optimal $\epsilon$ parameter value was guided by inspection of the 4 nearest neighbor (4-NN) distance plot (**Supplementary Figure S5A**). The optimal $\epsilon$ value was defined as the 4-NN value preceding the steep increase of the 4-NN distance.[12]

For visualization of assignment of the analysis subjects to three identified subsets ($HAP^{neg}$, $HAP^{int}$, $HAP^{high}$, based on the count of hyposmia/anosmia phenotype symptoms),the distance matrix was subjected to principal component analysis (PCA, function *PCAproj()*, package *pcaPP*)[13] and the first two principal components were presented in the plot (**Supplementary Figure S5B**). The R tools for clustering analysis are available at https://github.com/PiotrTymoszuk/cluster_tools.

#### Uni-variate modeling

Correlation of the candidate factors (**Supplementary Table S5**) with the sum of acute COVID-19 symptoms, presence of persistent COVID-19 symptoms, symptom relapse after the initial resolution and over 50% physical performance loss following COVID-19 was assessed with a series of univariate generalized linear models (symptom sum: assumed Poisson distribution of residuals, log-link functon; remaining responses: logistic regression) was utilized. To account for the sex and age bias as compared with the general population of COVID-19 convalescents, frequency weights were implemented in the modeling procedure based on the publicly available age and sex distributions of the COVID-19 convalescent populations in Tyrol (https://covid19-dashboard.ages.at/dashboard.html, access on 13thˆ July 2021) and Italy (https://www.epicentro.iss.it/coronavirus/bollettino/Bollettino-sorveglianza-integrata-COVID-19_7-luglio-2021.pdf, access on 13$^{th}$ July 2021) for the Tyrol and South Tyrol study cohorts, respectively (**Supplementary Table S6**. To address possible recall bias caused by retrospective surveying of the acute COVID-19 course features, continuous numeric observation time variable (test to survey compltetion time) was inceluded in every model as a confounder.

Significance of model estimates was determined by Wald Z test and p values were corrected for multiple comparisons with Benjamini-Hochberg method. Model quality and assumptions were checked by visual inspection of the residual plots. The tools used for serial modeling and model quality check are available at https://github.com/PiotrTymoszuk/lm_qc_tools. To obtain pooled $\beta$ estimates and their errors for the Tyrol and South Tyrol cohorts presented in the article text, the inverse variance method (function *metagen()*, package *meta*) was applied.[14,15]

The numbers of significant factors shared between the responses and cohorts were visualized as an upset plot.[16]

#### LASSO modeling

Identification of the most influential, independent factors associated with the presence of persistent COVID-19 symptoms, symptom relapse after the initial resolution and over 50% physical performance loss following COVID-19 was accomplished with LASSO (least absolute shrinkage and selection operator) 10-fold cross-validated generalized linear modeling (symptom sum responses: assumed Poisson distribution of residuals, log-link functon; remaining responses: logistic regression, function *cv.glmnet()* from *glmnet* package).[17,18]

The initial pool of variables used in the LASSO model construction based largely on the candidate variables for univariate modeling (**Supplementary Table S5**); however, two variables: 'Quality of support' and 'Contact to onset' were excluded due to over 30% missingness. To account for age and sex bias in the study populations, frequency weighting based was implemented in the models analogically to univariate modeling (**Supplementary Table S6**).

Model parameters were obtained for the lambda parameter set to 'lambda.1se' resulting in the output models with optimal regularization. Regression estimates of model co-variates with non-zero LASSO regression coeffcients were further analyzed and visualized. Measures of model fitness and error were derived from the cross-validated models using *assess.glmnet()* function, pseudo $R^2$ was calculated with the formula $pseudo - R^2 = 1 - deviance/null\ deviance$. Quality of model fit and assumptions were checked by visual inspection of the residual plots. The tools used for serial modeling and model quality check are available at https://github.com/PiotrTymoszuk/lasso_tools.

##### Prediction of development of persistent symptoms, symptom relapse and major physical performance impairment following COVID-19

To assess the ability of the LASSO models trained in the Tyrol cohort to predict development of persistent COVID-19 symptoms, symptom relapse and over 50% physical performance impairment following COVID-19 in the South Tyrol cohort, linear predictor scores (LPS) based on the models developed in the training cohort were calculated and the LPS prediction quality was assessed by receiver-operator characteristic (ROC) analysis with the functions from *Optimal.Cutpoints* and *plotROC* packages. The optimal LPS cutpoints were determined by Youden criterion. For LPS formulas, see: **Supplementary Table S11**.

### Data availability

As this study is still ongoing, the data will be made available on a serious request and made publicly available after the completion. The entire R analysis pipeline is available at https://github.com/PiotrTymoszuk/health-after-COVID19-analysis-pipeline.

#### Supplementary Tables

Table S1: **Variables queried directly and determined based on survey answers.**
Variable name: full variable name, Variable short name: short variable name used for plot labeling, Unit: variable unit, Description: variable description, Cutpoints: cutoffs used for variable stratification, Levels: variable strata, Variable type: survey module.
The table is available online.

Table S2: **Stratification of the acute symptom sum variables for modeling tasks**

| Cohort | Quartile | # acute symptoms | # acute NIP symptoms | # acute MOP symptoms |
|---|---|---|---|---|
| North Tyrol | Q1 | (0,9] | (0,6] | (0,1] |
| | Q2 | (9,13] | (6,8] | (1,3] |
| | Q3 | (13,18] | (8,10] | (3,5] |
| | Q4 | (18, 44] | (10, 12] | (5, 26] |
| South Tyrol | Q1 | (0,7] | (0,6] | (0,1] |
| | Q2 | (7,13] | (6,8] | (1,3] |
| | Q3 | (13,18] | (8,10] | (3,6] |
| | Q4 | (18, 44] | (10, 12] | (6, 26] |

Table S3: **Whole-cohort symptom prevalence and time changes of COVID-19 symptoms**
Symptom frequency at the given time point (first two, two to four and four weeks or longer after symptom onset) was determined as a percent of the entire cohort. Statistical significance of the time change in symptom frequency was assessed by $\chi^2$ test for trend and p values corrected for multiple comparisons with Benjamini-Hochberg method.
Time point: time interval after symptom onset, N: number of cohort members with the symptom.
The table is available online.

Table S4: **Assignment of acute and persistent COVID-19 symptoms to the phenotypes.**.

| Symptom character | Phenotype | Symptoms |
|---|---|---|
| acute, first two weeks | Multi-Organ Phenotype (MOP) | Wet cough, Dyspnea, Tachycardia, Palpitations, Abdominal pain, Nausea, Vomiting, Diarrhea, Confusion, Tingling feet, Tingling hands, Burning feet, Burning hands, Numb feet, Numb hands, Imp. walk, Imp. fine motor skills, Sleeplessness, Forgetfulness, Epilepsy, Swelling, Blue fingers/toes, Urticaria, Blistering rash, Blue marmorated skin, Red eyes |
| | Non-specific Inferion Phenotype (NIP) | Fever, Fatigue, Dry cough, Tachypnea, Joint pain, Muscle pain, Dim. appetite, Headache, Hypo/anosmia, Hypo/ageusia, Tiredness at day, Imp. concentration |
| persistent, more than four weeks | Fatigue Phenotype (FAP) | Fatigue, Tiredness at day, Imp. concentration, Forgetfulness |
| | Hyposmia/Anosmia Phenotype (HAP) | Hypo/anosmia, Hypo/ageusia |
| | Multi-Organ Phenotype (MOP) | Fever, Shivering, Sore throat, Running nose, Dry cough, Wet cough, Tachypnea, Dyspnea, Chest pain, Tachycardia, Palpitations, Joint pain, Bone pain, Muscle pain, Abdominal pain, Nausea, Vomiting, Dim. appetite, Diarrhea, Dizziness, Headache, Confusion, Tingling feet, Tingling hands, Burning feet, Burning hands, Numb feet, Numb hands, Imp. walk, Imp. fine motor skills, Sleeplessness, Epilepsy, Swelling, Blue fingers/toes, Urticaria, Blistering rash, Blue marmorated skin, Red eyes |

Table S5: **Candidate variables in modeling tasks.**.

| Response | Method | Co-variates |
|---|---|---|
| Symptom relapse | logistic regression | Responder, Sex, Education, Region, Age, Household size, #CoV in household, BMI before CoV, Empl. status, Empl. sector, Surv. completion, Autoimmunity, Anemia, Hypertension, Pre-CoV depr/anxiety, Diabetes, Pre-CoV epilepsy, Dementia, Surg. Before CoV, Freq. resp. inf., CVD, Allergy, Malignancy, Paresthesia, GI disease, Lung dis., Freq. bact. Inf, MS, Sleep apnea, Bruxism, CKD, Parkinson, Pre-CoV sleep disord., Stroke, Embolism, Transplantation, # comorb., Daily medic., Daily immunsupp., Daily corticost., Daily ACE, Daily analgesia, Daily anticoag., Flu vacc., Pneumo. vacc, Smoking, Inf. contact, Contact-onset, Quarantine, Auth. support, Supp. quality, CoV outbreak, Hair loss, Weight loss, Subj. CoV percept., Contact physician, CoV no therapy, CoV anti-pyretic, CoV antibiotic, Acute fever, Acute shivering, Acute sore throat, Acute running nose, Acute fatigue, Acute dry cough, Acute wet cough, Acute tachypnea, Acute dyspnea, Acute chest pain, Acute tachycardia, Acute palpitations, Acute joint pain, Acute bone pain, Acute muscle pain, Acute abd. pain, Acute nausea, Acute vomiting, Acute dim. appetite, Acute diarrhea, Acute dizziness, Acute headache, Acute hyposmia/anosmia, Acute hypogeusia/ageusia, Acute confusion, Acute tingling feet, Acute tingling hands, Acute burning feet, Acute burning hands, Acute numb feet, Acute numb hands, Acute imp. walk, Acute imp. f. m. s., Acute sleeplessness, Acute tiredness at day, Acute imp. concentration, Acute forgetfulness, Acute fever, Acute swelling, Acute blue fingers/toes, Acute urticaria, Acute blistering rash, Acute bl. marm. skin., Acute red eyes, Obs.time, # acute symptoms, # acute NIP symptoms, # acute MOP symptoms |
| Sum of acute symptoms | GLM Poisson | Responder, Sex, Education, Region, Age, Household size, #CoV in household, BMI before CoV, Empl. status, Empl. sector, Surv. completion, Autoimmunity, Anemia, Hypertension, Pre-CoV depr/anxiety, Diabetes, Pre-CoV epilepsy, Dementia, Surg. Before CoV, Freq. resp. inf., CVD, Allergy, Malignancy, Paresthesia, GI disease, Lung dis., Freq. bact. Inf, MS, Sleep apnea, Bruxism, CKD, Parkinson, Pre-CoV sleep disord., Stroke, Embolism, Transplantation, # comorb., Daily medic., Daily immunsupp., Daily corticost., Daily ACE, Daily analgesia, Daily anticoag., Flu vacc., Pneumo. vacc, Smoking, Inf. contact, Contact-onset, Quarantine, Auth. support, Supp. quality, CoV outbreak, Contact physician, CoV no therapy, CoV anti-pyretic, CoV antibiotic, Obs.time |

Table S5: **Candidate variables in modeling tasks.**. Continued

| Response | Method | Co-variates |
|---|---|---|
| Presence of persistent COVID-19 symptoms | logistic regression | Responder, Sex, Education, Region, Age, Household size, #CoV in household, BMI before CoV, Empl. status, Empl. sector, Surv. completion, Autoimmunity, Anemia, Hypertension, Pre-CoV depr/anxiety, Diabetes, Pre-CoV epilepsy, Dementia, Surg. Before CoV, Freq. resp. inf., CVD, Allergy, Malignancy, Paresthesia, GI disease, Lung dis., Freq. bact. Inf, MS, Sleep apnea, Bruxism, CKD, Parkinson, Pre-CoV sleep disord., Stroke, Embolism, Transplantation, # comorb., Daily medic., Daily immunsupp., Daily corticost., Daily ACE, Daily analgesia, Daily anticoag., Flu vacc., Pneumo. vacc, Smoking, Inf. contact, Contact-onset, Quarantine, Auth. support, Supp. quality, CoV outbreak, Hair loss, Weight loss, Subj. CoV percept., Contact physician, CoV no therapy, CoV anti-pyretic, CoV antibiotic, Acute fever, Acute shivering, Acute sore throat, Acute running nose, Acute fatigue, Acute dry cough, Acute wet cough, Acute tachypnea, Acute dyspnea, Acute chest pain, Acute tachycardia, Acute palpitations, Acute joint pain, Acute bone pain, Acute muscle pain, Acute abd. pain, Acute nausea, Acute vomiting, Acute dim. appetite, Acute diarrhea, Acute dizziness, Acute headache, Acute hyposmia/anosmia, Acute hypogeusia/ageusia, Acute confusion, Acute tingling feet, Acute tingling hands, Acute burning feet, Acute burning hands, Acute numb feet, Acute numb hands, Acute imp. walk, Acute imp. f. m. s., Acute sleeplessness, Acute tiredness at day, Acute imp. concentration, Acute forgetfulness, Acute fever, Acute swelling, Acute blue fingers/toes, Acute urticaria, Acute blistering rash, Acute bl. marm. skin., Acute red eyes, Obs.time, # acute symptoms, # acute NIP symptoms, # acute MOP symptoms |
| Over 50% physical performance loss | logistic regression | Responder, Sex, Education, Region, Age, Household size, #CoV in household, BMI before CoV, Empl. status, Empl. sector, Surv. completion, Autoimmunity, Anemia, Hypertension, Pre-CoV depr/anxiety, Diabetes, Pre-CoV epilepsy, Dementia, Surg. Before CoV, Freq. resp. inf., CVD, Allergy, Malignancy, Paresthesia, GI disease, Lung dis., Freq. bact. Inf, MS, Sleep apnea, Bruxism, CKD, Parkinson, Pre-CoV sleep disord., Stroke, Embolism, Transplantation, # comorb., Daily medic., Daily immunsupp., Daily corticost., Daily ACE, Daily analgesia, Daily anticoag., Flu vacc., Pneumo. vacc, Smoking, Inf. contact, Contact-onset, Quarantine, Auth. support, Supp. quality, CoV outbreak, Asymptomatic, Hair loss, Weight loss, Subj. CoV percept., Contact physician, CoV no therapy, CoV anti-pyretic, CoV antibiotic, Acute fever, Acute shivering, Acute sore throat, Acute running nose, Acute fatigue, Acute dry cough, Acute wet cough, Acute tachypnea, Acute dyspnea, Acute chest pain, Acute tachycardia, Acute palpitations, Acute joint pain, Acute bone pain, Acute muscle pain, Acute abd. pain, Acute nausea, Acute vomiting, Acute dim. appetite, Acute diarrhea, Acute dizziness, Acute headache, Acute hyposmia/anosmia, Acute hypogeusia/ageusia, Acute confusion, Acute tingling feet, Acute tingling hands, Acute burning feet, Acute burning hands, Acute numb feet, Acute numb hands, Acute imp. walk, Acute imp. f. m. s., Acute sleeplessness, Acute tiredness at day, Acute imp. concentration, Acute forgetfulness, Acute fever, Acute swelling, Acute blue fingers/toes, Acute urticaria, Acute blistering rash, Acute bl. marm. skin., Acute red eyes, Obs.time, # acute symptoms, # acute NIP symptoms, # acute MOP symptoms |

Table S6: **Weights applied to age and strata in modeling tasks.**.
Frequency weights for each age and sex strata were based on the age and sex distribution of the general COVID-19 convalescent populations in Tyrol and Italy, for the Tyrol and South Tyrol study cohorts, respectively.
Convalescents: number of convalescents in the given strata in the Tyrol or Italy population.

| Cohort | Age strata | Sex | Convalescents | Freq. weight |
|---|---|---|---|---|
| North Tyrol | <5 | male | 492 | 0.0078720 |
| | <5 | female | 402 | 0.0064320 |
| | 5-14 | male | 2750 | 0.0440000 |
| | 5-14 | female | 2574 | 0.0411840 |
| | 15-24 | male | 4846 | 0.0775360 |
| | 15-24 | female | 4478 | 0.0716480 |
| | 25-34 | male | 5241 | 0.0838560 |
| | 25-34 | female | 4960 | 0.0793600 |
| | 35-44 | male | 4411 | 0.0705760 |
| | 35-44 | female | 4711 | 0.0753760 |
| | 45-54 | male | 5209 | 0.0833440 |
| | 45-54 | female | 5658 | 0.0905280 |
| | 55-64 | male | 4467 | 0.0714720 |
| | 55-64 | female | 4053 | 0.0648480 |
| | 65-74 | male | 1875 | 0.0300000 |
| | 65-74 | female | 2000 | 0.0320000 |
| | 75-84 | male | 1294 | 0.0207040 |
| | 75-84 | female | 1597 | 0.0255520 |
| | >84 | male | 474 | 0.0075840 |
| | >84 | female | 1008 | 0.0161280 |
| South Tyrol | 0-9 | male | 120929 | 0.0293421 |
| | 10-19 | male | 214102 | 0.0519495 |
| | 20-29 | male | 256298 | 0.0621879 |
| | 30-39 | male | 258715 | 0.0627743 |
| | 40-49 | male | 324116 | 0.0786431 |
| | 50-59 | male | 358095 | 0.0868878 |
| | 60-69 | male | 233361 | 0.0566225 |
| | 70-79 | male | 151073 | 0.0366562 |
| | 80-89 | male | 76247 | 0.0185005 |
| | Over 90 | male | 12787 | 0.0031026 |
| | 0-9 | female | 112727 | 0.0273520 |
| | 10-19 | female | 196809 | 0.0477535 |
| | 20-29 | female | 248432 | 0.0602793 |
| | 30-39 | female | 271502 | 0.0658769 |
| | 40-49 | female | 356740 | 0.0865590 |
| | 50-59 | female | 373925 | 0.0907287 |
| | 60-69 | female | 221200 | 0.0536717 |
| | 70-79 | female | 157086 | 0.0381152 |
| | 80-89 | female | 126781 | 0.0307620 |
| | Over 90 | female | 50426 | 0.0122353 |

Table S7: **Results of univariate modeling.**
Correlation of candidate factors with the sum of acute COVID-19 symptoms (first two weeks after symptom onset), presence of persistent symptoms (four weeks or longer after the symptom onset), symptom relapse after the initial resolution and over 50% physical performance loss following COVID-19 was investigated with a series of sex- and age-weighted ordinary generalized linear models (GLM) with inclusion of continuous observation time variable (test to completion interval) as a confounder. Estimate significances were obtained with Wald Z test and p values corrected for multiple comparisons with Benjamini-Hochberg method.
The table is available online.

Table S8: **Pooled univariate $\beta$ estimates of the univariate acute symptom modeling for the risk factors presented in the text.**
Pooled estimates of the univariate Poisson regression of the acute symptom number in the Tyrol and South Tyrol cohort were calculated with the inverse variance method. P values were corrected for multiple comparisons with Benjamini-Hochberg method.

| Co-variate | Exp. estimate | 2.5% CI | 97.5% CI | Significance |
|---|---|---|---|---|
| BMI before COVID-19: obesity | 1.210 | 1.140 | 1.290 | p = 2e-230 |
| BMI before COVID-19: overweigth | 1.070 | 1.010 | 1.130 | p = 2.2e-276 |
| Pre-CoV depression/anxiety: yes | 1.260 | 1.160 | 1.360 | p = 1.6e-131 |
| Freq. resp. infections: yes | 1.330 | 1.220 | 1.450 | p = 3e-112 |
| Pre-CoV sleep disorders: yes | 1.330 | 1.220 | 1.440 | p = 1.3e-122 |
| Pulmonary disease: yes | 1.300 | 1.180 | 1.420 | p = 3e-95 |
| Sum of co-morbidities: 1 | 1.060 | 0.997 | 1.120 | p = 4.6e-263 |
| Sum of co-morbidities: 2 | 1.170 | 1.090 | 1.250 | p = 1.5e-188 |
| Sum of co-morbidities: 3 and more | 1.470 | 1.380 | 1.550 | p = 2.9e-233 |
| Contact with physician: yes | 1.550 | 1.500 | 1.600 | p = 0 |
| Sex: male | 0.767 | 0.711 | 0.823 | p = 1.7e-157 |
| COVID-19 anti-pyretic therapy: yes | 1.370 | 1.320 | 1.430 | p = 0 |
| COVID-19 no therapy: yes | 0.709 | 0.657 | 0.761 | p = 8.3e-156 |
| COVID-19 antibiotic therapy: yes | 1.450 | 1.370 | 1.520 | p = 3.5e-276 |

Table S9: **Pooled univariate $\beta$ estimates of the univariate long COVID feature modeling for the risk factors presented in the text.**
Pooled estimates of the univariate logistic regression in the Tyrol and South Tyrol cohort were calculated with the inverse variance method. P values were corrected for multiple comparisons with Benjamini-Hochberg method.

| Co-variate | Response | Exp. estimate | 2.5% CI | 97.5% CI | Significance |
|---|---|---|---|---|---|
| Acute multi-organ phenotype symptoms: 4Q | Over 50% physical performance loss | 22.30 | 21.50 | 23.00 | p = 0 |
| | Presence of persistent COVID-19 symptoms | 8.69 | 8.39 | 8.99 | p = 0 |
| | Symptom relapse | 7.39 | 7.08 | 7.69 | p = 0 |
| Confusion acute COVID-19: yes | Over 50% physical performance loss | 4.32 | 3.97 | 4.67 | p = 5.2e-130 |
| | Presence of persistent COVID-19 symptoms | 5.33 | 5.06 | 5.59 | p = 0 |
| | Symptom relapse | 3.74 | 3.51 | 3.98 | p = 5.8e-209 |
| Dizziness acute COVID-19: yes | Over 50% physical performance loss | 3.28 | 2.93 | 3.64 | p = 4.4e-74 |
| | Presence of persistent COVID-19 symptoms | 3.42 | 3.23 | 3.62 | p = 9.1e-256 |
| | Symptom relapse | 2.82 | 2.62 | 3.02 | p = 8.2e-165 |
| Dyspnea acute COVID-19: yes | Over 50% physical performance loss | 4.92 | 4.57 | 5.27 | p = 1.2e-169 |
| | Presence of persistent COVID-19 symptoms | 3.12 | 2.91 | 3.32 | p = 4.7e-190 |
| | Symptom relapse | 2.62 | 2.41 | 2.83 | p = 1.3e-131 |
| Fatigue acute COVID-19: yes | Over 50% physical performance loss | 7.25 | 6.13 | 8.37 | p = 8.8e-37 |
| | Presence of persistent COVID-19 symptoms | 6.43 | 6.08 | 6.78 | p = 6.1e-281 |
| | Symptom relapse | 9.88 | 9.25 | 10.50 | p = 5.7e-209 |
| Forgetfulness acute COVID-19: yes | Over 50% physical performance loss | 4.54 | 4.20 | 4.88 | p = 6.8e-148 |
| | Presence of persistent COVID-19 symptoms | 4.88 | 4.67 | 5.09 | p = 0 |
| | Symptom relapse | 3.15 | 2.94 | 3.36 | p = 5.9e-195 |
| Palpitations acute COVID-19: yes | Over 50% physical performance loss | 3.43 | 3.07 | 3.80 | p = 8.3e-76 |
| | Presence of persistent COVID-19 symptoms | 3.83 | 3.55 | 4.12 | p = 5.3e-152 |
| | Symptom relapse | 3.96 | 3.70 | 4.22 | p = 7.1e-198 |
| | Over 50% physical performance loss | 22.40 | 21.60 | 23.20 | p = 0 |

Table S9: **Pooled univariate $\beta$ estimates of the univariate long COVID feature modeling for the risk factors presented in the text.**
Pooled estimates of the univariate logistic regression in the Tyrol and South Tyrol cohort were calculated with the inverse variance method. P values were corrected for multiple comparisons with Benjamini-Hochberg method. *(continued)*

| Co-variate | Response | Exp. estimate | 2.5% CI | 97.5% CI | Significance |
|---|---|---|---|---|---|
| Sum of acute symptoms: 4Q | Presence of persistent COVID-19 symptoms | 14.90 | 14.60 | 15.20 | p = 0 |
| | Symptom relapse | 10.00 | 9.67 | 10.30 | p = 0 |
| Tachycardia acute COVID-19: yes | Over 50% physical performance loss | 4.28 | 3.95 | 4.62 | p = 8.4e-137 |
| | Presence of persistent COVID-19 symptoms | 3.57 | 3.36 | 3.79 | p = 3e-231 |
| | Symptom relapse | 3.31 | 3.10 | 3.52 | p = 1.8e-204 |
| Tachypnea acute COVID-19: yes | Over 50% physical performance loss | 6.05 | 5.61 | 6.50 | p = 3.9e-156 |
| | Presence of persistent COVID-19 symptoms | 3.63 | 3.45 | 3.82 | p = 0 |
| | Symptom relapse | 2.53 | 2.32 | 2.73 | p = 2.9e-126 |

Table S10: **Results of LASSO modeling.**
To identify independent factors correlating with the sum of acute COVID-19 symptoms (first two weeks after symptom onset), presence and sum of persistent symptoms (four weeks or longer after the symptom onset), symptom relapse after the initial resolution and over 50% physical performance loss following COVID-19 was sex- and age-weighted LASSO generalized linear modeling was applied. Model co-variates with non-zero regression coefficients are presented.
The table is available online.

Table S11: **Formulas of linear predictor scores developed with LASSO modeling in the Tyrol training cohort.**

| Response | Score formula |
| --- | --- |
| Symptom relapse | LPS = -1.08 × (Intercept) + 0.129 × (# acute symptoms: 4Q) + 0.253 × (# acute MOP symptoms: 4Q) + 0.234 × (Contact physician: yes) + 0.249 × (Acute chest pain: yes) + 0.14 × (Acute imp. concentration: yes) + 0.126 × (Acute forgetfulness: yes) |
| Presence of persistent COVID-19 symptoms | LPS = -1.29 × (Intercept) + -0.183 × (Daily ACE: yes) + 0.133 × (# acute MOP symptoms: 4Q) + 0.338 × (Surv. completion: winter spring 2021) + 0.0654 × (Hair loss: yes) + 0.214 × (Contact physician: yes) + 0.322 × (Acute tachypnea: yes) + 0.275 × (Acute dyspnea: yes) + 0.104 × (Acute palpitations: yes) + 0.00902 × (Acute dizziness: yes) + 0.404 × (Acute hyposmia/anosmia: yes) + 0.0621 × (Acute hypogeusia/ageusia: yes) + 0.214 × (Acute sleeplessness: yes) + 0.191 × (Acute imp. concentration: yes) + 0.629 × (Acute forgetfulness: yes) + 0.132 × (Obs.time: 61 - 120 days) + 0.375 × (Obs.time: 121 - 180 days) + 0.347 × (Obs.time: more than 180 days) |
| Over 50% physical performance loss | LPS = -2.82 × (Intercept) + 0.23 × (# acute symptoms: 4Q) + 0.319 × (# acute MOP symptoms: 4Q) + 0.403 × (Pre-CoV depr/anxiety: yes) + 0.142 × (Contact physician: yes) + 0.028 × (Acute tachypnea: yes) + 0.0203 × (Acute chest pain: yes) + 0.00996 × (Acute confusion: yes) + 0.0571 × (Acute tingling feet: yes) + 0.0726 × (Acute forgetfulness: yes) |

### Supplementary Figures

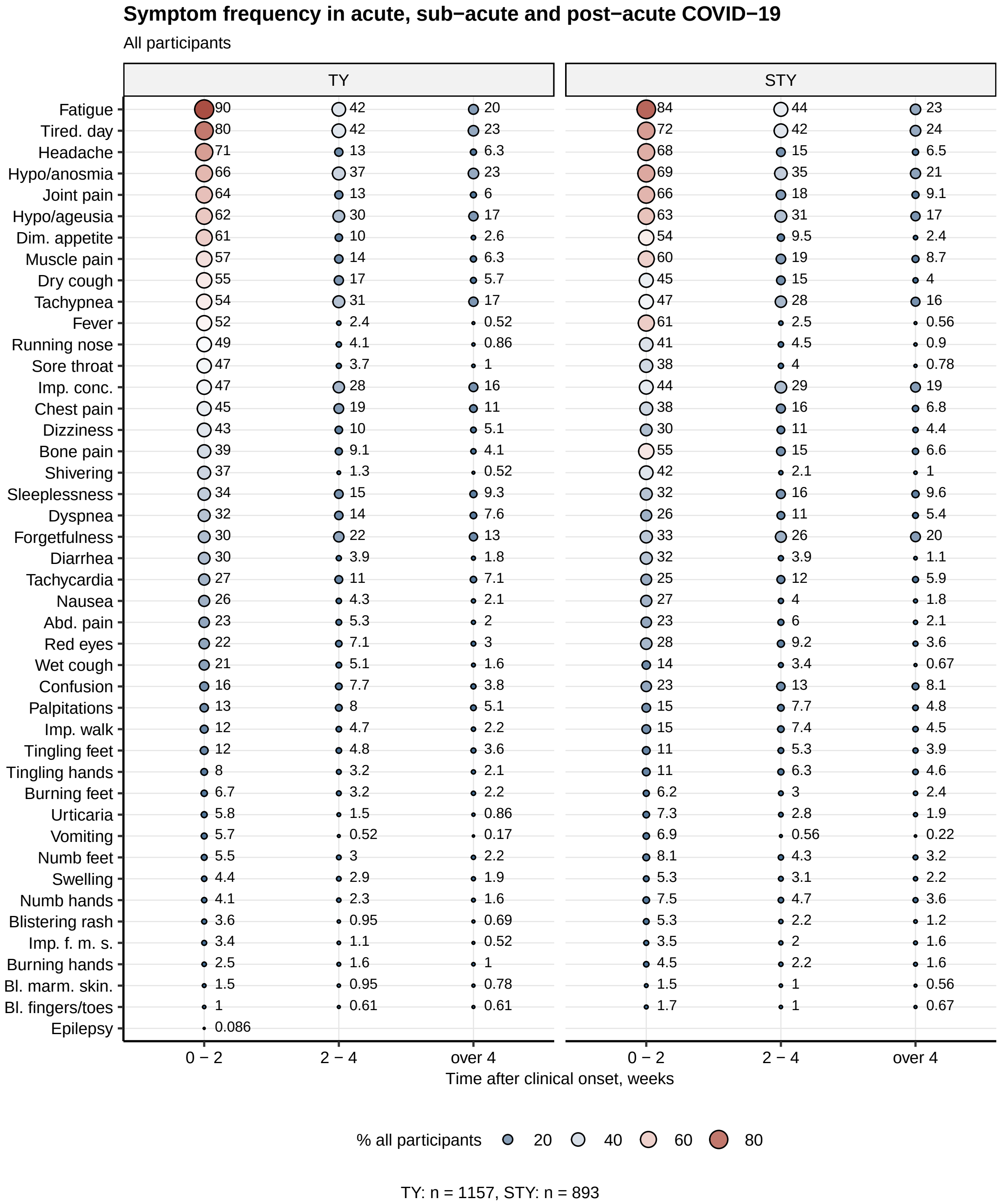

Figure S1: Frequency of acute, sub-acute and persistent symptoms of COVID-19 in the entire study cohorts.

**Supplementary Figure S1. Frequency of acute, sub-acute and persistent symptoms of COVID-19 in the entire study cohorts.**

Symptom frequencies in the first two, two to four weeks and four weeks or longer after symptom onset were calculated as of the entire cohort. % within the entire cohort are presented as a bubble plot. Point size and color represents the percentage. Numbers of complete observations are indicated below the plot.

Tired. day: tiredness at day, imp. conc.: impaired concentration, abd. pain: abdominal pain, imp. walk: impaired walk, dim. appetite = diminished appetite, imp.f.m.s: impaired fine motor skills, TY: Tyrol, STY: South Tyrol cohort.

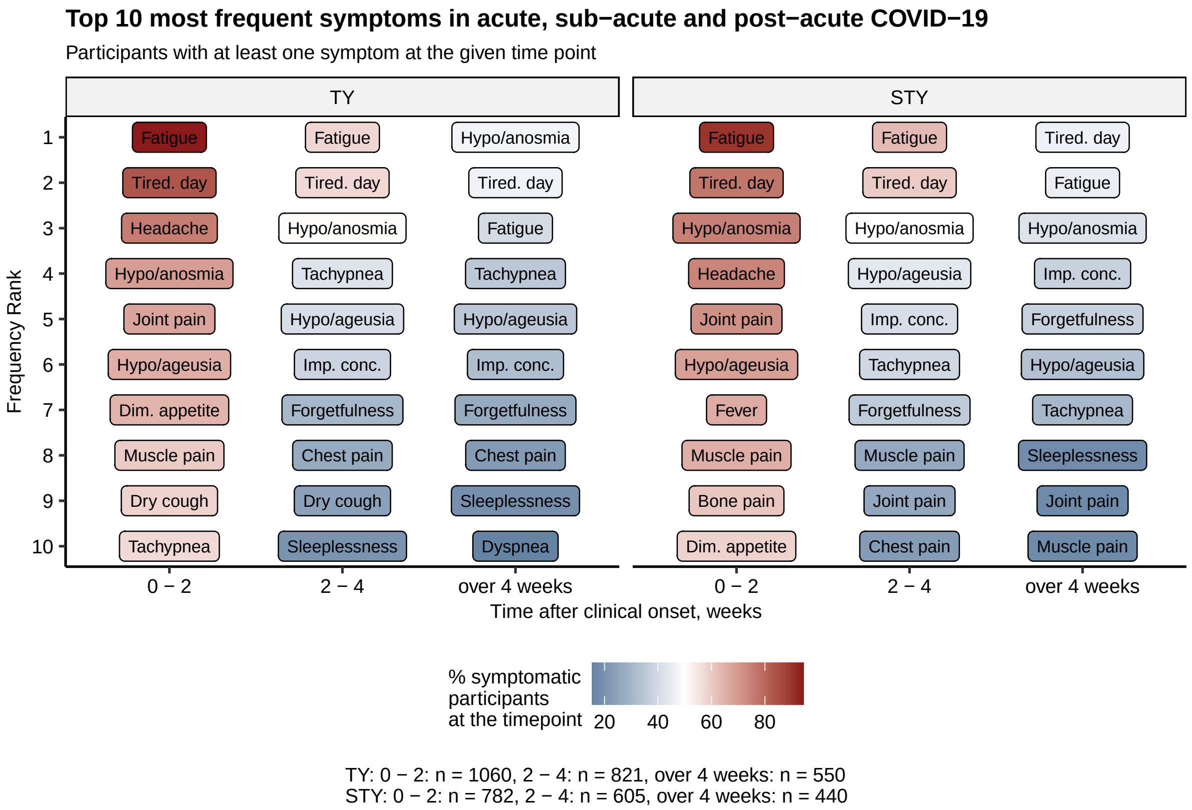

Figure S2: Ten most frequent acute, sub-acute and persistent symptoms of COVID-19.

**Supplementary Figure S2. Ten most frequent acute, sub-acute and persistent symptoms of COVID-19.**

Symptom frequencies were calculated as percentages of the individuals reporting at least one symptom in the first two, two to four weeks and four weeks or longer after symptom onset. Ten most frequent symptoms in the symptomatic subset are presented. Numbers of complete observations are indicated below the plot.

Tired. day: tiredness at day, imp. conc.: impaired concentration, dim. appetite = diminished appetite, TY: Tyrol, South: STY cohort.

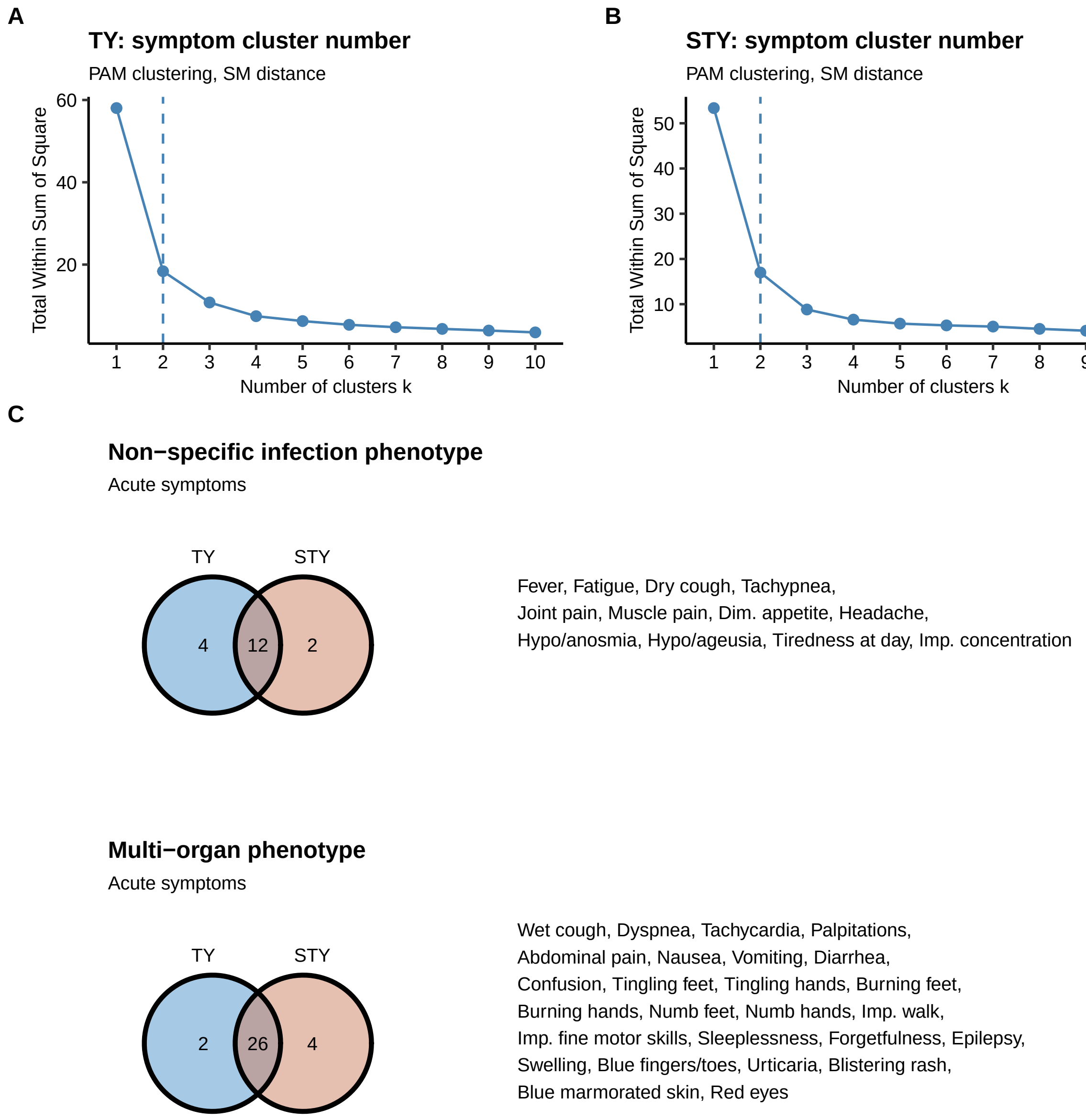

Figure S3: Definition of acute COVID-19 symptom phenotypes.

**Supplementary Figure S3. Definition of symptom phenotypes in acute COVID-19.**

Simple matching distances between the symptoms of COVID-19 present in the first two weeks after symptom onset were subjected to clustering with PAM (partitioning around medoids) algorithm. Clusters of the same symptoms in both cohorts were termed consensus phenotypes.

**(A)** Plots of total within-cluster sum of squares as a function of cluster number used to guide the decision on the optimal cluster count by the 'curve elbow' method. Dashed vertical lines represent the chosen numbers of acute symptom clusters.

**(B)** Numbers of symptoms assigned to clusters in each study cohort and common consensus clusters termed the non-specific infection phenotype (NIP) and the multi-organ phenotype (MOP) presented in Venn plots. Consensus symptoms assigned to each phenotype are listed next to the plot.

Tired. day: tiredness at day, imp. conc.: impaired concentration, abd. pain: abdominal pain, imp. walk: impaired walk, dim. appetite: diminished appetite, imp.f.m.s: impaired fine motor skills, TY: Tyrol, STY: South Tyrol cohort.

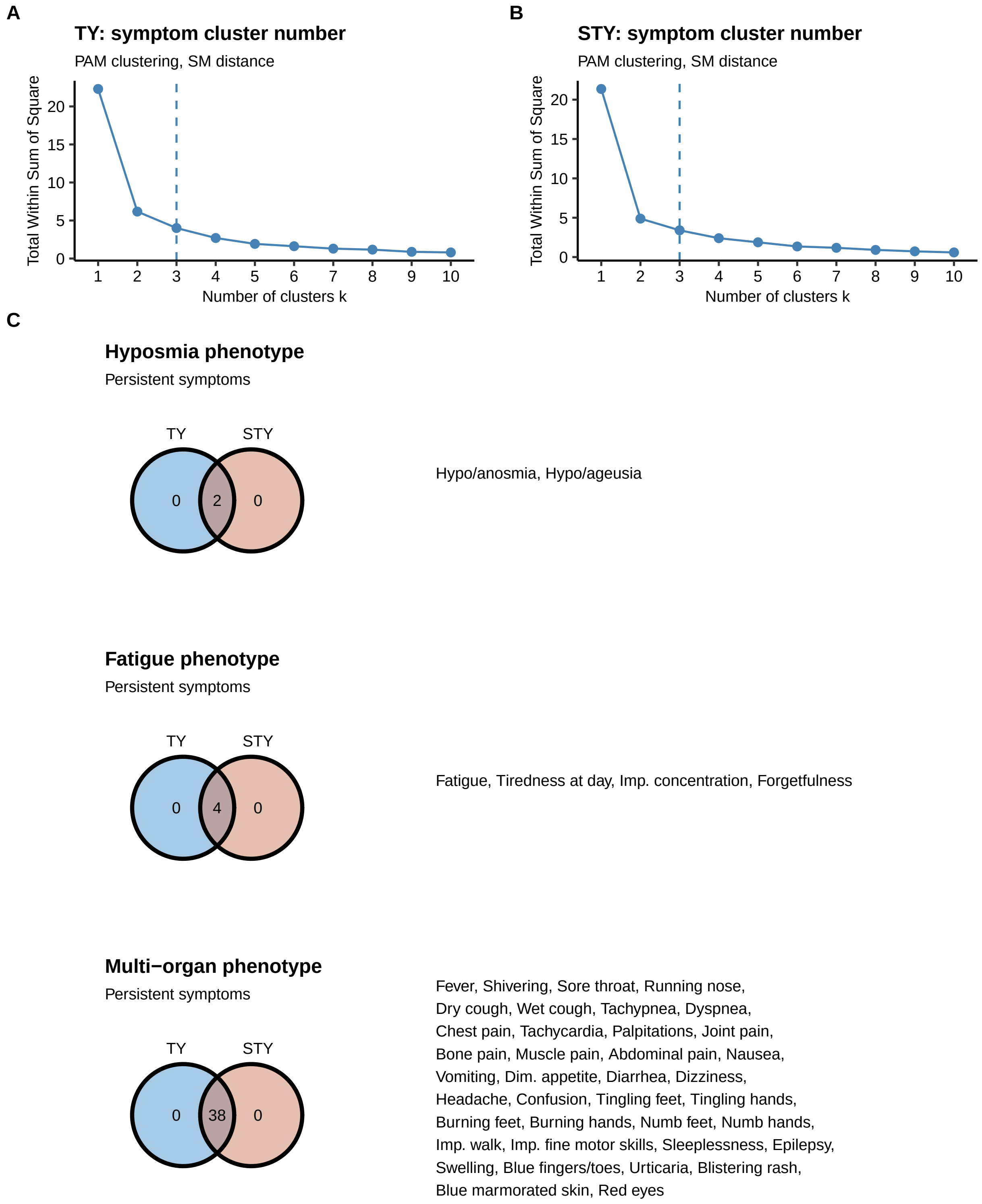

Figure S4: Definition of persistent COVID-19 symptom phenotypes.

**Supplementary Figure S4. Definition of persistent COVID-19 symptom phenotypes.**
Simple matching distances between the symptoms of COVID-19 present for four weeks or longer symptom

onset were subjected to clustering with PAM (partitioning around medoids) algorithm. Clusters of the same symptoms in both cohorts were termed consensus phenotypes.

**(A)** Plots of total within-cluster sum of squares as a function of cluster number used to guide the decision on the optimal cluster count by the 'curve elbow' method. Dashed vertical lines represent the chosen numbers of persistent symptom clusters.

**(B)** Numbers of symptoms assigned to clusters in each study cohort and common consensus clusters termed the hyposmia/anosmia phenotype (HAP), fatigue phenotype (FAP) and the multi-organ phenotype (MOP) presented in Venn plots. Consensus symptoms assigned to each phenotype are listed next to the plot.

Tired. day: tiredness at day, imp. conc.: impaired concentration, abd. pain: abdominal pain, imp. walk: impaired walk, dim. appetite: diminished appetite, imp.f.m.s: impaired fine motor skills. TY: Tyrol, STY: South Tyrol cohort.

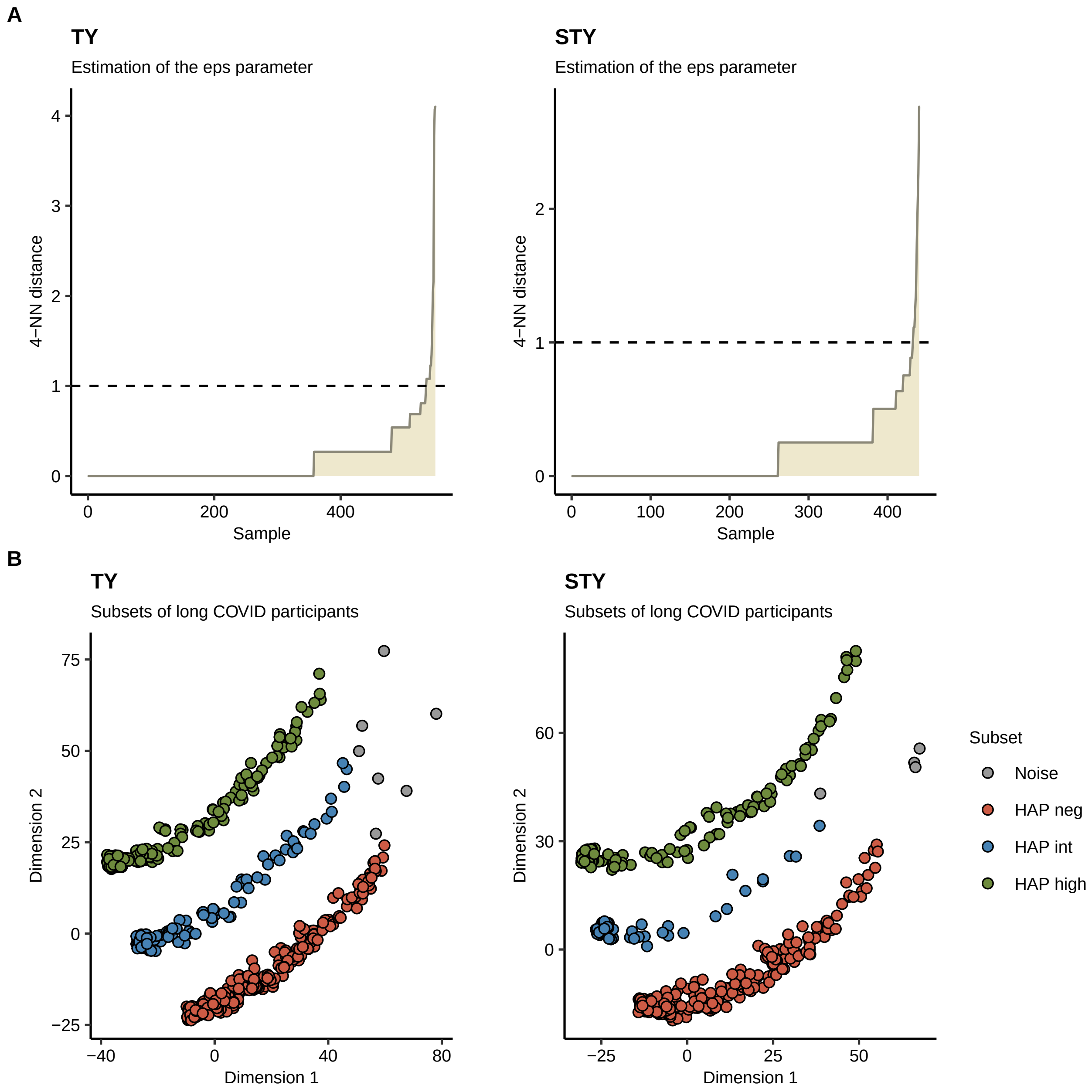

Figure S5: Definition of subsets of participants suffering from persistent COVID-19 symptoms.

**Supplementary Figure S5. Definition of subsets of participants suffering from persistent COVID-19 symptoms.**

The subset of the study cohorts suffering from COVID-19 symptoms lasting for four weeks or longer after symptom onset was classified into three subsets: $HAP^{neg}$, $HAP^{int}$ and $HAP^{high}$ in regard to the count of the persistent hyposmia/anosmia (HAP), fatigue (FAP) and multi-organ phenotype symptoms (MOP) (**Supplementary Figure S4**) by the DBSCAN clustering procedure with Manhattan distance.

**(A)** Plots of the sorted 4-nearest neighbor (4-NN) Manhattan distances as a function of the observation number used to guide the decision on the optimal value of the $\epsilon$. The optimal $\epsilon$ value was defined as the 4-NN value preceding the steep increase of the 4-NN distance.

**(B)** Assignment of the participants with persistent COVID-19 symptoms to the $HAP^{neg}$, $HAP^{int}$ and $HAP^{high}$ subsets. The plots display two first principal components derived from principal component analysis (PCA) of the Manhattan distance matrix.

Tyrol: $HAP^{neg}$ n = 268, $HAP^{int}$ n = 102, $HAP^{high}$ n = 173. South Tyrol: $HAP^{neg}$ n = 242, $HAP^{int}$ n = 58, $HAP^{high}$ n = 136.

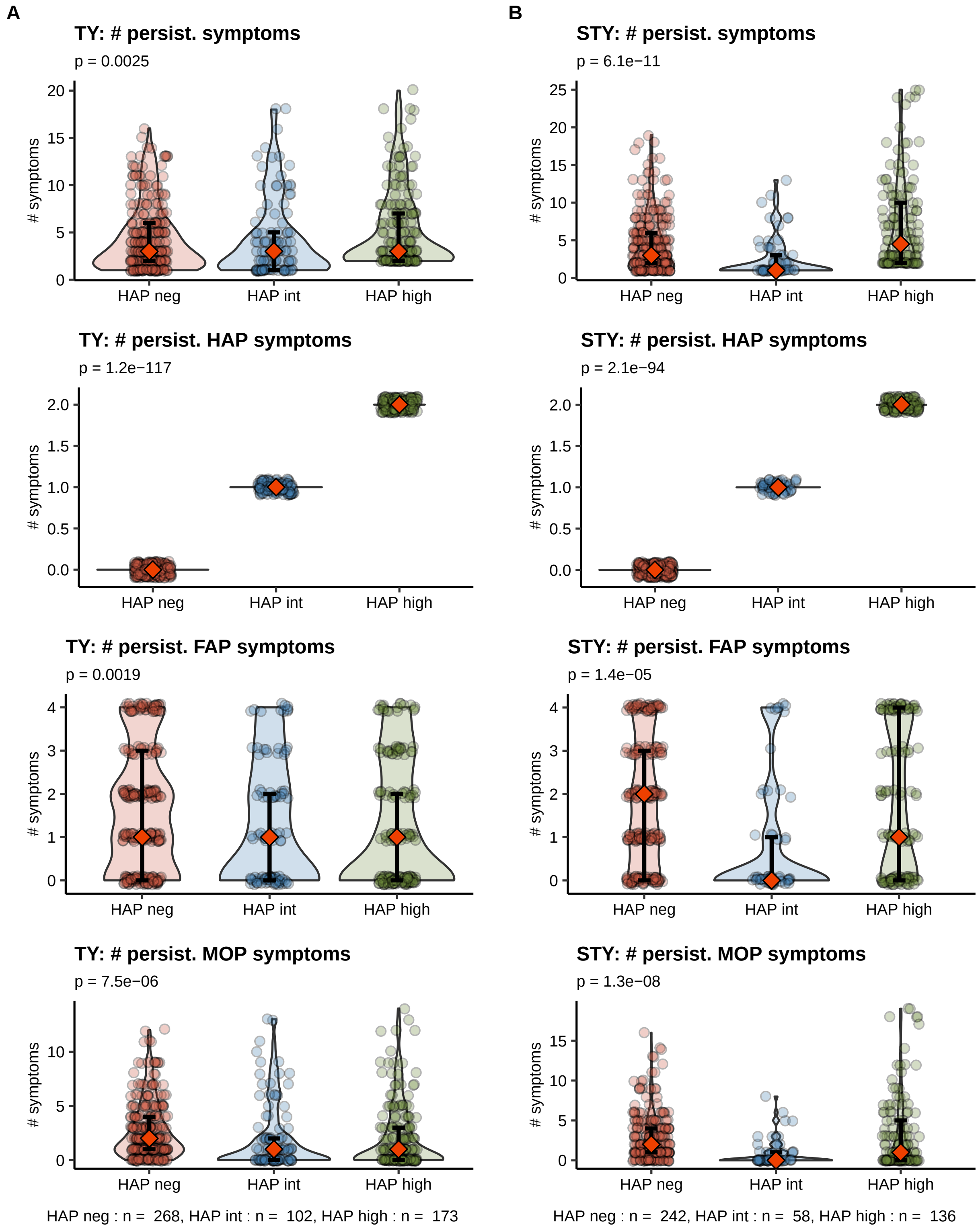

Figure S6: Sums of all, hyposmia/anosmia, fatigue and multi-organ phenotype symptoms in participants with persistent COVID-19 complaints.

**Supplementary Figure S6. Sums of all, hyposmia/anosmia, fatigue and multi-organ phenotype symptoms in participants with persistent COVID-19 complaints.**

Study participants suffering from persistent COVID-19 complaints for four weeks or longer after symptom onset were assigned to the $HAP^{neg}$, $HAP^{int}$ and $HAP^{high}$ subsets (**Supplementary Figure S5**) defined by the counts of the hyposmia/anosmia (HAP) fatigue (FAP) and multi-organ phenotype symptoms (**Supplementary Figure S4**).

Sums (#) of all hyposmia, fatigue, and multi-organ phenotype symptoms in the Tyrol **(A)** and South Tyrol **(B)** cohort presented as violin plots. Each point denotes a single participant, orange diamonds and whiskers represent medians with IQR. Statistical significance was assessed by Kruskal-Wallis test, p values corrected for multiple comparisons with Benjamini-Hochberg method are presented in plot headings. Numbers of complete observations are indicated under the bottom plots.

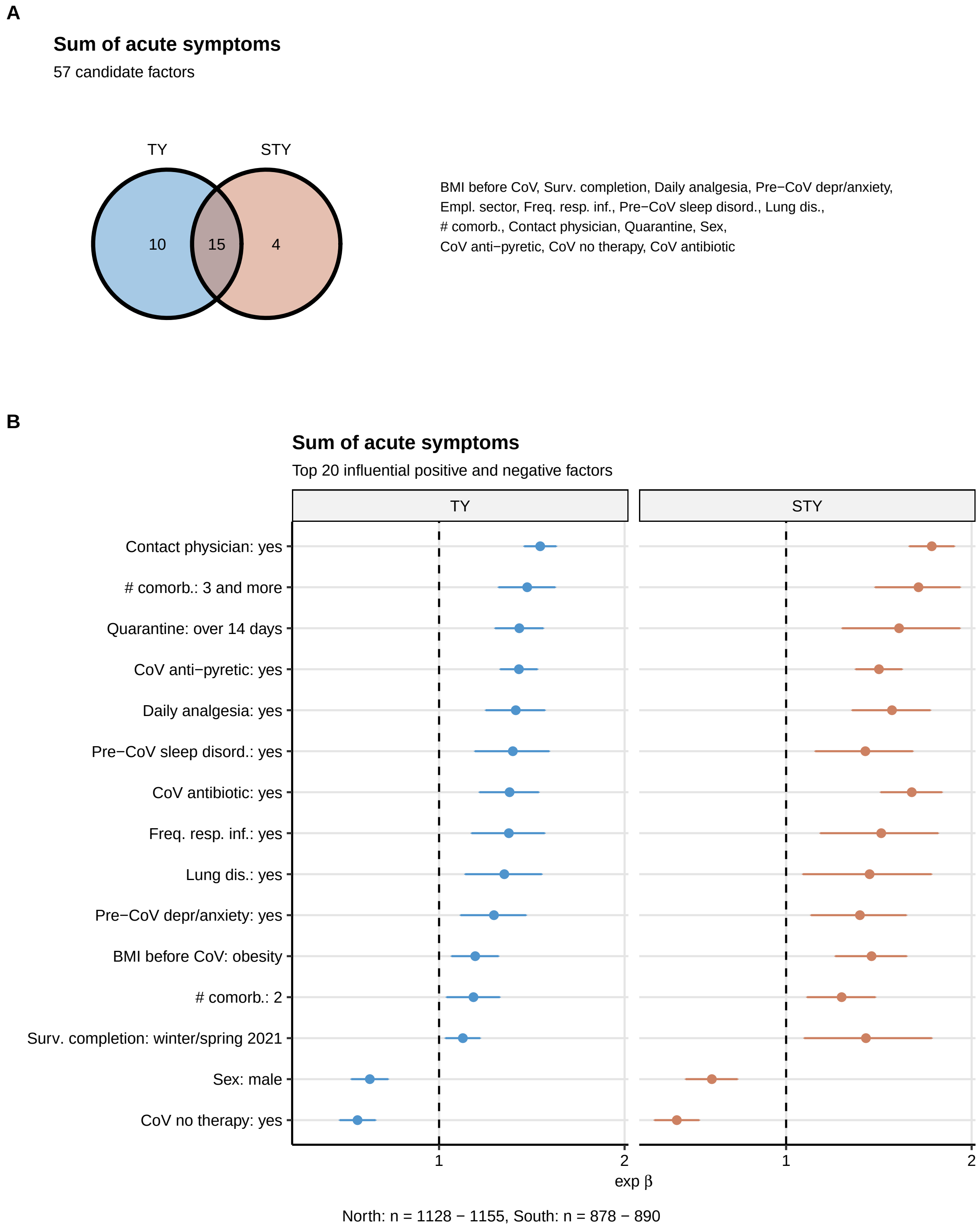

Figure S7: Identification of factors associated with the acute COVID-19 symptom sum by univariate modeling.

**Supplementary Figure S7. Identification of factors associated with the acute COVID-19 symptom sum by univariate modeling.**

Correlation of 55 candidate factors with the sum of acute COVID-19 symptoms (first two weeks after symptom onset) was investigated with a series of sex- and age-weighted ordinary generalized linear models (GLM, assumed Poisson distribution of residuals, log link function) with inclusion of continuous observation time variable (test to completion interval) as a confounder. Estimate significance was determined by Wald Z-test and corrected for multiple comparisons with Benjamini-Hochberg method. For full list of significant factors, see: **Supplementary Table S7**.

**(A)** Numbers of significant factors associated with the acute symptom burden in each of the study populations presented in a Venn plot. Common significant factors are listed next to the plot.

**(B)** Values of regression coefficients of the top 20 most influential significant factors correlating with the acute symptom burden in the Tyrol cohort and verified in the South Tyrol cohort presented in a Forest plot. Points represent $\beta$ estimate values, whiskers represent the 95% confidence interval. Numbers of complete observations are presented below the plot.

Surv. completion: survey completion, Daily analgesia: daily analgetic medication before COVID-19, Pre-CoV depr/anxiety,: depression or anxiety before COVID-19, Empl. sector: employment sector, Freq. resp. inf.: frequent respiratory infections before COVID-19, Pre-CoV sleep disord.: sleep disorders before COVID-19, Lung dis.: lung disease before COVID-19, # comorb.: number of pre-existing co-morbidities, CoV antibiotic/CoV anti-pyretic/CoV no therapy: antibiotic/anti-pyretic/no symptom-specific treatment during acute COVID-19, Contact physician: contact with a physician during quarantine.

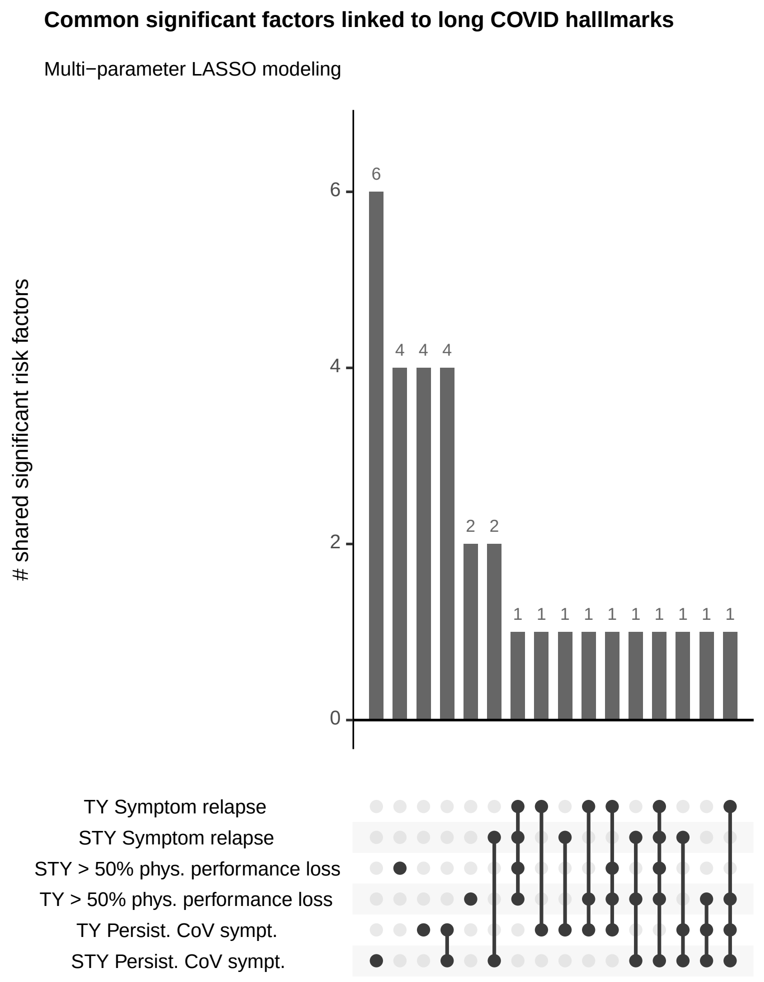

Figure S8: Common significant co-variates of symptom persistence, relapse and >50% physical performance loss following COVID-19.

**Supplementary Figure S8. Common significant co-variates of symptom persistence, relapse and >50% physical performance loss following COVID-19.**

Correlation of candidate factors with development of persistent COVID-19 symptoms (four weeks or longer after symptom onset), symptom relapse or >50% physical performance loss was investigated with a series of sex- and age-weighted ordinary logistic models with inclusion of continuous observation time variable (test to completion interval) as a confounder. Estimate significance was determined by Wald Z-test and corrected for multiple comparisons with Benjamini-Hochberg method. Numbers of significant co-variates shared between the readouts and cohorts are visualized in the bar/upset plot. Factors significant for all investigated readouts in both study cohorts are listed next to the plot in the alphabetical order.

MOP: multi-organ phenotype, NIP: non-specific infection phenotype, #: number of, abd.: abdominal, imp.: impaired, subj. CoV percept.: subjective perception of acute COVID-19.

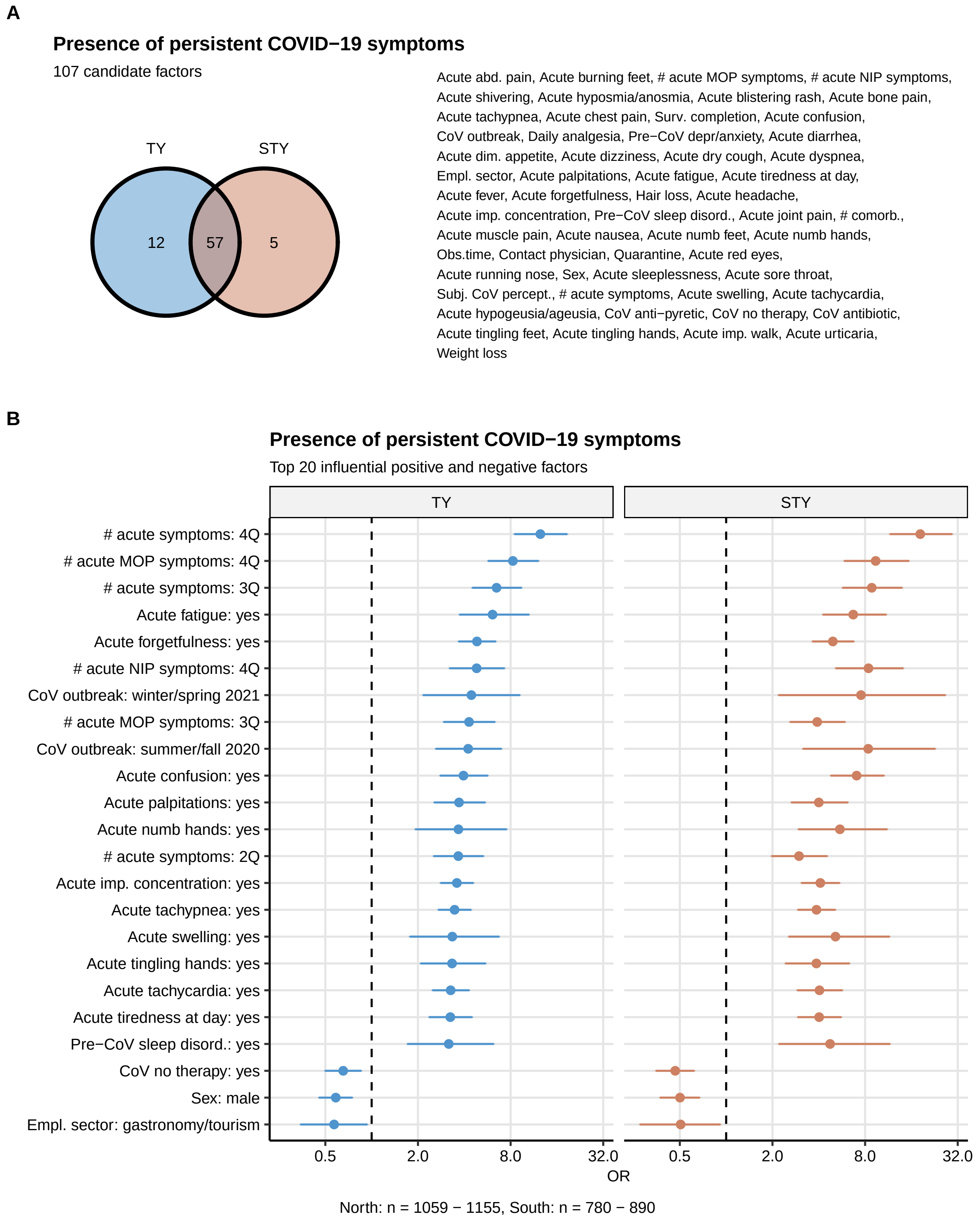

Figure S9: Identification of factors associated with development of persistent COVID-19 symptoms.

**Supplementary Figure S9. Identification of factors associated with development of persistent COVID-19 symptoms.**

Correlation of 105 candidate factors with development of persistent COVID-19 symptoms (four weeks or longer after symptom onset) was investigated with a series of sex- and age-weighted ordinary logistic models with inclusion of continuous observation time variable (test to completion interval) as a confounder. Estimate significance was determined by Wald Z-test and corrected for multiple comparisons with Benjamini-Hochberg method. For full list of significant factors, see: **Supplementary Table S7**.

**(A)** Numbers of significant factors associated with development of persistent COVID-19 symptoms in each of the study populations presented in a Venn plot. Common significant factors are listed next to the plot.

**(B)** Values of odds ratios (OR) of the top 20 most influential significant factors correlating with development of persistent COVID-19 symptoms in the Tyrol cohort and verified in the South Tyrol cohort presented in a Forest plot. Points represent OR estimate values, whiskers represent the 95% confidence interval. Numbers of complete observations are presented below the plot.

Empl. sector: employment sector, # acute MOP/NIP symptoms: number of acute multi-organ/non-specific infection phenotype symptoms, Surv. completion: survey completion, Obs. time: test to study completion time interval, Pre-CoV depr/anxiety,: depression or anxiety before COVID-19, Freq. resp. inf: frequent respiratory infections, # comorb.: number of pre-existing co-morbidities, CoV antibiotic/CoV anti-pyretic/CoV no therapy: antibiotic/anti-pyretic/no symptom-specific treatment during acute COVID-19, Contact physician: contact with a physician during quarantine, imp. walk: impaired walk, # acute symptoms: total number of acute symptoms, 2Q, 3Q, 4Q: second, third, fourth quartile of symptom number.

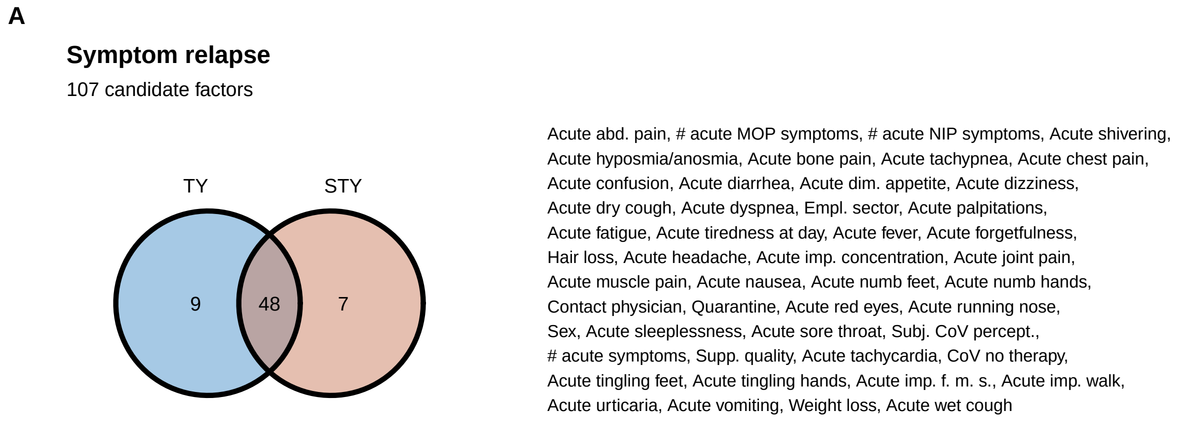

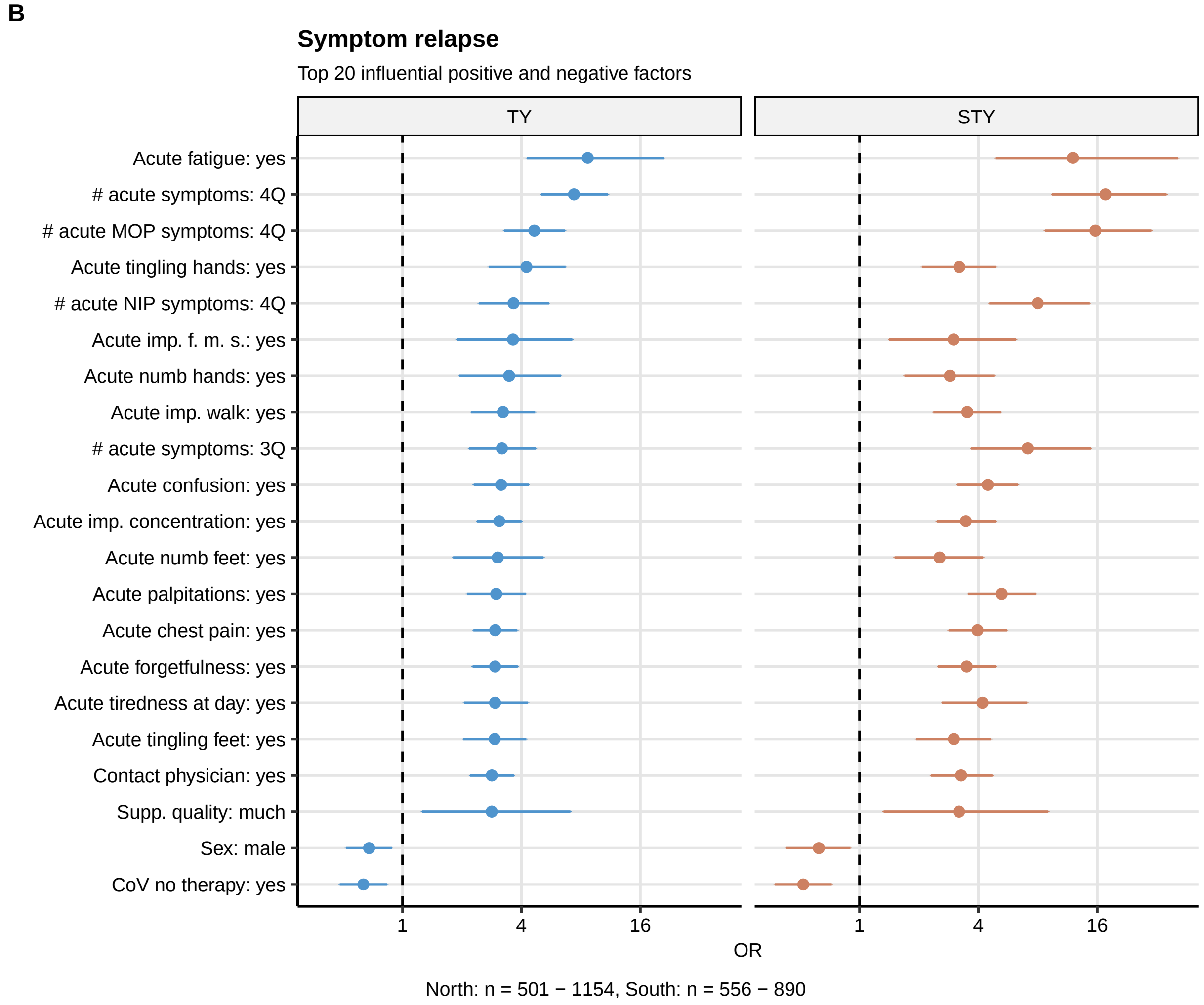

Figure S10: Identification of factors associated symptom relapse after the initial resolution.

**Supplementary Figure S10. Identification of factors associated symptom relapse after the initial resolution.**

Correlation of 105 candidate factors with symptom relapse after the initial resolution was investigated with a series of sex- and age-weighted ordinary logistic models with inclusion of continuous observation time variable (test to completion interval) as a confounder. Estimate significance was determined by Wald Z-test and corrected for multiple comparisons with Benjamini-Hochberg method. For full list of significant factors, see: **Supplementary Table S7**.

**(A)** Numbers of significant factors associated with symptom relapse in each of the study populations presented in a Venn plot. Common significant factors are listed next to the plot.

**(B)** Values of odds ratios (OR) of the top 20 most influential significant factors correlating with relapse in the Tyrol cohort and verified in the South Tyrol cohort presented in a Forest plot. Points represent OR estimate values, whiskers represent the 95% confidence interval. Numbers of complete observations are presented below the plot.

abd. pain: abdominal pain, dim. appetite: diminished appetite, Subj. CoV percept: subjective perception of acute COVID-19, Supp. quality: quality of authority support during quarantine, imp. f. m. s.: impaired fine motor skills, imp.walk: impaired walk, # acute MOP/NIP symptoms: number of acute multi-organ/non-specific infection phenotype symptoms, # acute symptoms: total number of acute symptoms, 2Q, 3Q, 4Q: second, third, fourth quartile of symptom number.

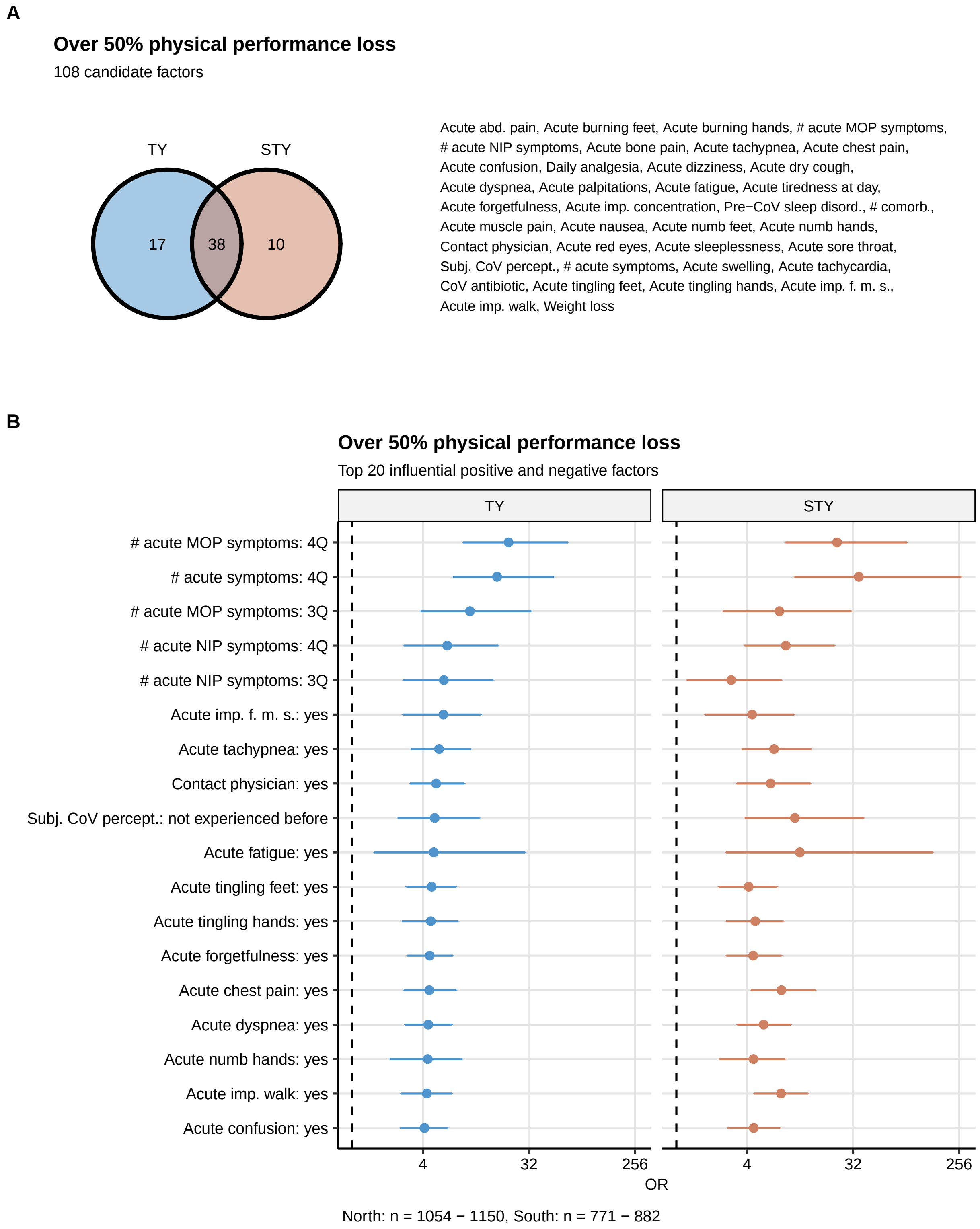

Figure S11: Identification of factors associated with major physical performance impairment following COVID-19.

**Supplementary Figure S10. Identification of factors associated with major physical performance impairment following COVID-19.**

Correlation of 106 candidate factors with over 50% physical performance impairment following COVID-19 was investigated with a series of sex- and age-weighted ordinary logistic models with inclusion of continuous observation time variable (test to completion interval) as a confounder. Estimate significance was determined by Wald Z-test and corrected for multiple comparisons with Benjamini-Hochberg method. For full list of significant factors, see: **Supplementary Table S7**.

**(A)** Numbers of significant factors associated with major physical performance impairment relapse in each of the study populations presented in a Venn plot. Common significant factors are listed next to the plot.

**(B)** Values of odds ratios (OR) of the top 20 most influential significant factors correlating with major physical performance impairment in the Tyrol cohort and verified in the South Tyrol cohort presented in a Forest plot. Points represent OR estimate values, whiskers represent the 95% confidence interval. Numbers of complete observations are presented below the plot.

abd. pain: abdominal pain, Pre-CoV sleep disord.: sleep disorders before COVID-19, # comorb.: number of pre-existing co-morbidities, Contact physician: contact with a physician during quarantine, Subj. CoV percept.: subjective perception of COVID-19, CoV antibiotic: symptom-specific antibiotic treatment during acute COVID-19, imp. f. m. s.: impaired fine motor skills, imp. walk: impaired walk, # acute MOP/NIP symptoms: number of acute multi-organ/non-specific infection phenotype symptoms, # acute symptoms: total number of acute symptoms, 2Q, 3Q, 4Q: second, third, fourth quartile of symptom number.

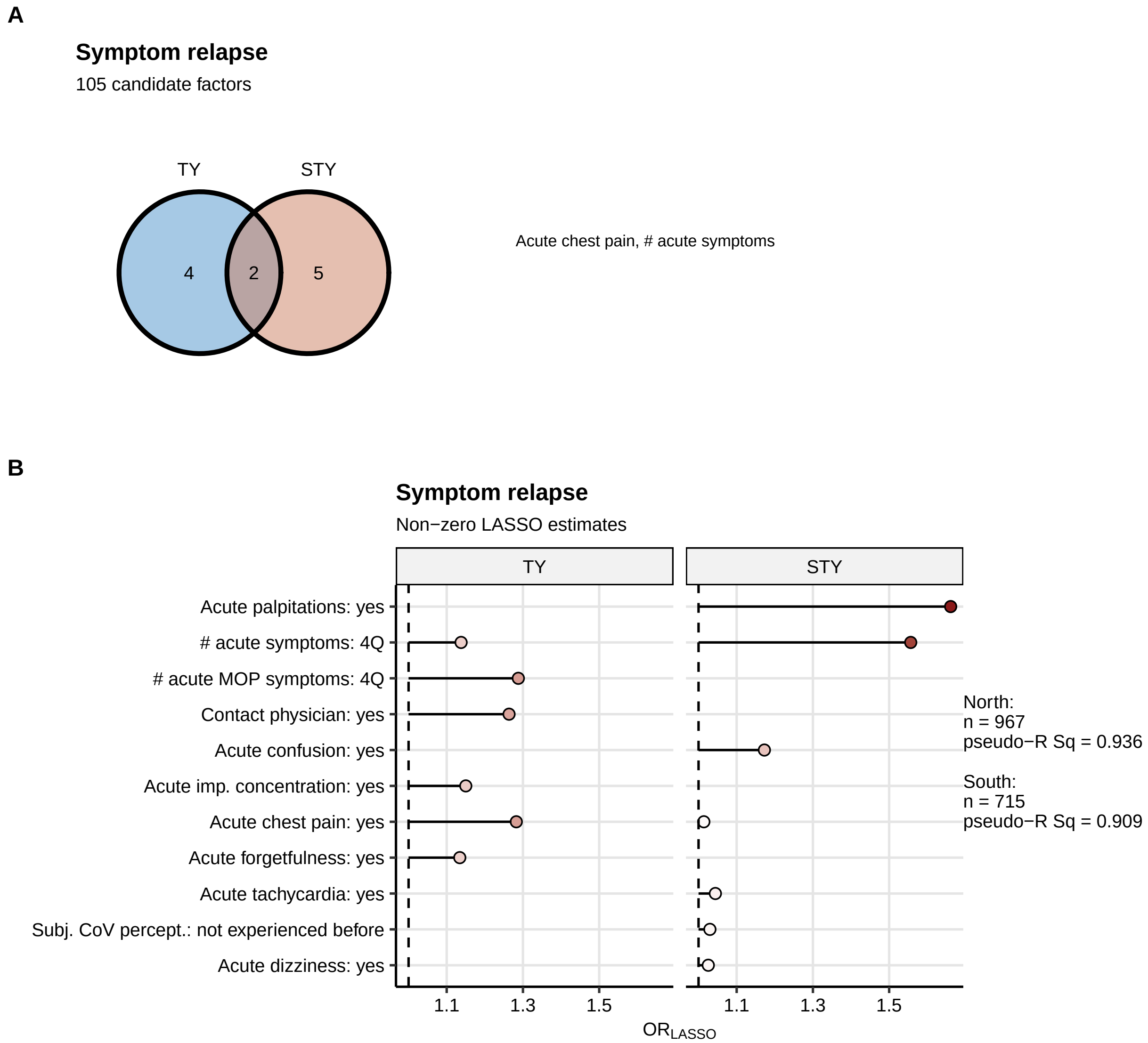

Figure S12: Identification of independent factors associated with symptom relapse after the initial resolution.___

**Supplementary Figure S12. Identification of independent factors associated with symptom relapse after the initial resolution.**

Correlation of 105 candidate factors with the risk of symptom relapse after the initial resolution was investigated by sex- and age-weighted LASSO (least absolute shrinkage and selection operator) generalized linear modeling (logistic regression) in the Tyrol and South Tyrol cohorts.

**(A)** Numbers of significant independent factors associated with symptom relapse in each of the study populations presented in a Venn plot. Common significant factors are listed next to the plot.

**(B)** Values of non-zero regression coefficents of the significant factors correlating with symptom relapse in the plot. Numbers of complete observations, mean absolute errors (MAE) and pseudo-$R^2$ statistics for each LASSO model are presented next to the plot.

imp. concentration: impaired concentration, Contact physician: contact with a physician during quarantine, # acute MOP symptoms: number of acute multi-organ phenotype symptoms, 2Q, 3Q, 4Q: second, third, fourth quartile of symptom number.

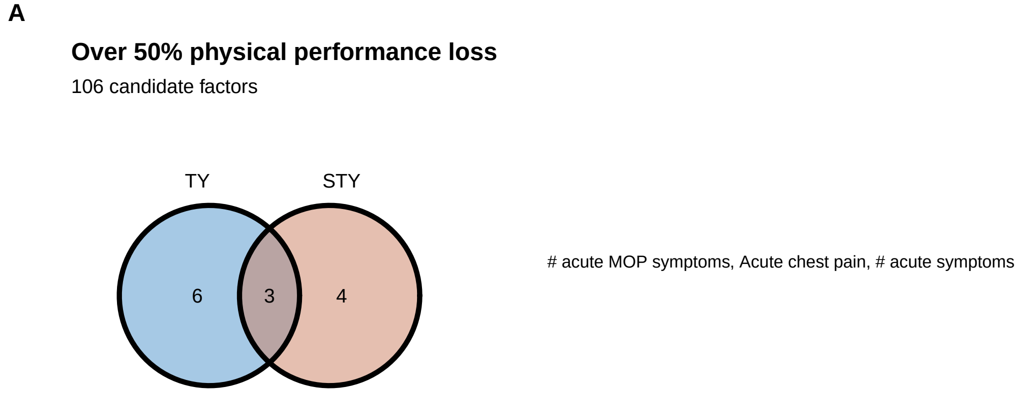

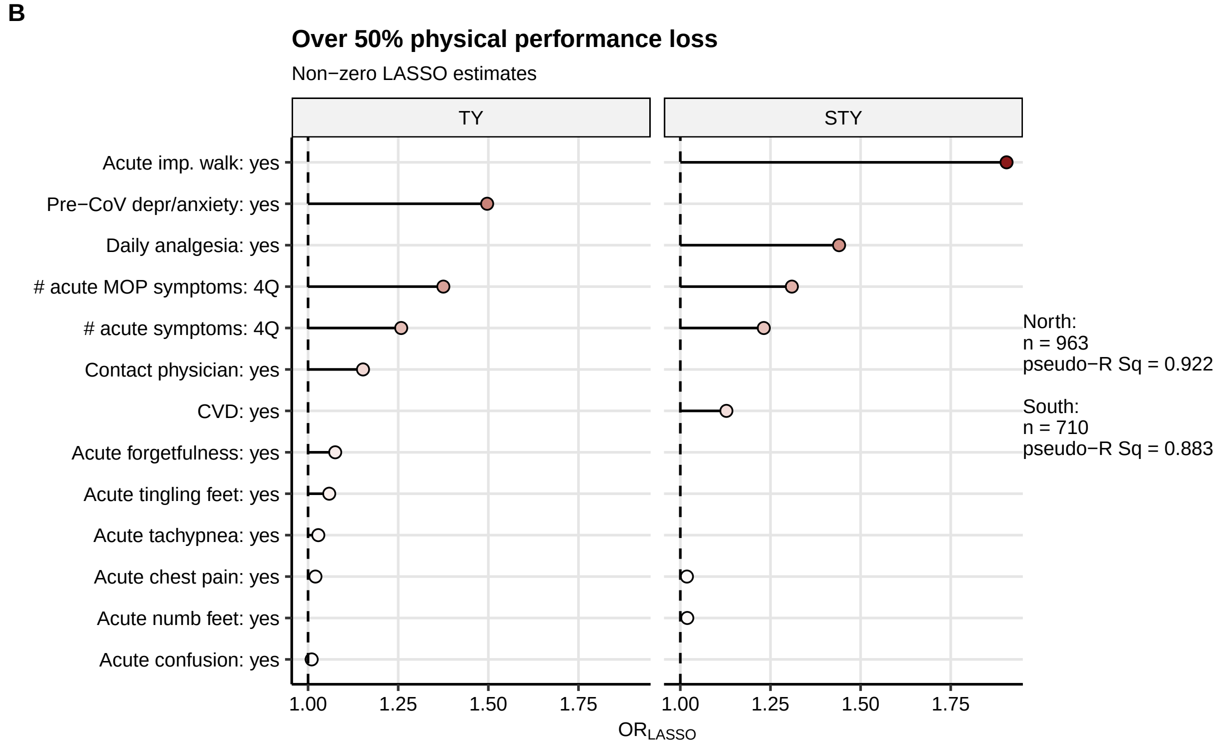

Figure S13: Identification of independent factors associated with major physical performance impairment following COVID-19.

**Supplementary Figure S13. Identification of independent factors associated with major physical performance impairment following COVID-19.**

Correlation of 106 candidate factors with over 50% physical performance impairment following COVID-19 was investigated by sex- and age-weighted LASSO (least absolute shrinkage and selection operator) generalized linear modeling (logistic regression) in the Tyrol and South Tyrol cohorts.

**(A)** Numbers of significant independent factors associated with major physical performance impairment in each of the study populations presented in a Venn plot. Common significant factors are listed next to the plot.

**(B)** Values of non-zero regression coefficents of the significant factors correlating with major physical performance impairment in the plot. Numbers of complete observations, mean absolute errors (MSE) and pseudo-$R^2$ statistics for each LASSO model are presented next to the plot.

Daily analgesia: daily analgetic medication before COVID-19, Pre-CoV depr/anxiety: depression or anxiety before COVID-19, Subj. CoV percept.: subjective perception of acute COVID-19, # acute MOP symptoms: number of acute multi-organ phenotype symptoms, # acute symptoms: total number of acute symptoms, 2Q, 3Q, 4Q: second, third, fourth quartile of symptom number.

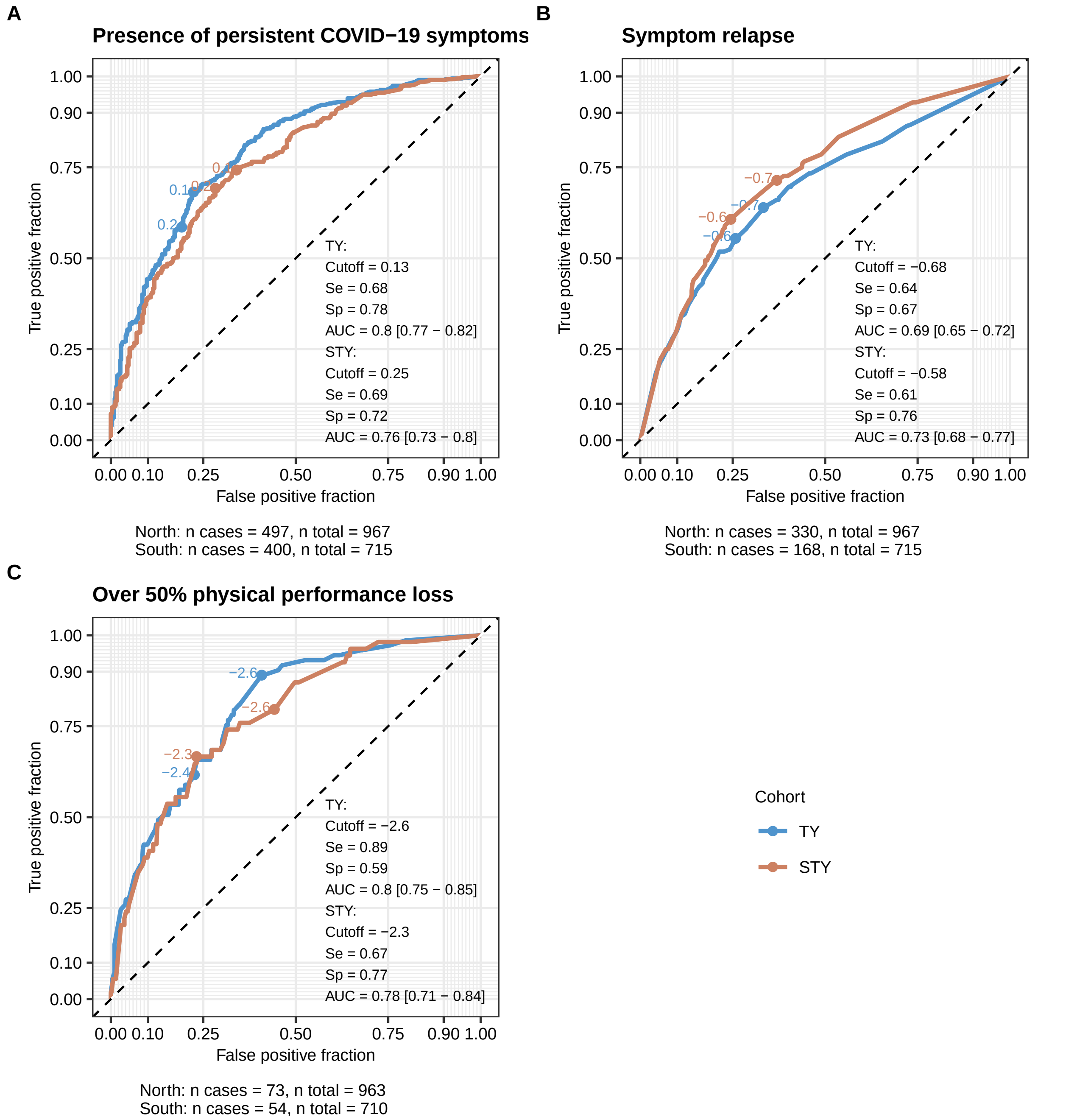

Figure S14: Prediction of persistent symtom development, symptom relapse and major physical performance loss by LASSO models trained in the Tyrol cohort.

**Figure S14. Prediction of persistent symtom development, symptom relapse and major physical performance loss by LASSO models trained in the Tyrol cohort.**

Correlation of multiple candidate features with the risk of developing persistent COVID-19 symptoms (four weeks or longer after symptom onset) **(A)**, symptom relapse after initial resolution **(B)** and over 50% physical performance loss following COVID-19 **(C)** was investigated with LASSO (least absolute shrinkage and selection operator) generalized linear modeling (logistic regression) in the Tyrol training cohort. Based on the trained models, linear predictor scores (LPS) were calculated for the training cohort and the test South Tyrol cohort. Prediction accuracy of the LPS in the both study collectives was investigated by receiver-operator characteristic (ROC) analysis.

Cutoff: optimal LPS cutoff, Se: sensitivity, Sp: specificity, AUC: area under the curve with 95% CI. Number of cases and the numbers of complete observations are indicated below the plots.
